# Acute COVID-19 severity and 16-month mental morbidity trajectories in patient populations of six nations

**DOI:** 10.1101/2021.12.13.21267368

**Authors:** Ingibjörg Magnúsdóttir, Anikó Lovik, Anna Bára Unnarsdóttir, Daniel McCartney, Helga Ask, Kadri Kõiv, Lea Arregui Nordahl Christoffersen, Sverre Urnes Johnson, Andrew McIntosh, Anna K. Kähler, Archie Campbell, Arna Hauksdóttir, Chloe Fawns-Ritchie, Christian Erikstrup, Dorte Helenius, Drew Altschul, Edda Bjork Thordardottir, Elías Eyþórsson, Emma M. Frans, Gunnar Tómasson, Harpa Lind Jónsdóttir, Harpa Rúnarsdóttir, Henrik Hjalgrim, Hrönn Harðardóttir, Juan González-Hijón, Karina Banasik, Khoa Manh Dinh, Li Lu, Lili Milani, Lill Trogstad, Maria Didriksen, Omid V. Ebrahimi, Patrick F. Sullivan, Per Minor Magnus, Qing Shen, Ragnar Nesvåg, Reedik Mägi, Runólfur Pálsson, Sisse Rye Ostrowski, Thomas Werge, Asle Hoffart, David J Porteous, Fang Fang, Jóhanna Jakobsdóttir, Kelli Lehto, Ole A Andreassen, Ole B. V. Pedersen, Thor Aspelund, Unnur Anna Valdimarsdóttir

## Abstract

**BACKGROUND:** The aim of this multinational study was to assess the development of adverse mental health symptoms among individuals diagnosed with COVID-19 in the general population by acute infection severity up to 16 months after diagnosis.

**METHODS:** Participants consisted of 247 249 individuals from seven cohorts across six countries (Denmark, Estonia, Iceland, Norway, Scotland, and Sweden) recruited from April 2020 through August 2021. We used multivariable Poisson regression to contrast symptom-prevalence of depression, anxiety, COVID-19 related distress, and poor sleep quality among individuals with and without a diagnosis of COVID-19 at entry to respective cohorts by time (0-16 months) from diagnosis. We also applied generalised estimating equations (GEE) analysis to test differences in repeated measures of mental health symptoms before and after COVID-19 diagnosis among individuals ever diagnosed with COVID-19 over time.

**FINDINGS:** A total of 9979 individuals (4%) were diagnosed with COVID-19 during the study period and presented overall with a higher symptom burden of depression (prevalence ratio [PR] 1·18, 95% confidence interval [95% CI] 1·03-1·36) and poorer sleep quality (1·13, 1·03-1·24) but not with higher levels of symptoms of anxiety or COVID-19 related distress compared with individuals without a COVID-19 diagnosis. While the prevalence of depression and COVID-19 related distress attenuated with time, the trajectories varied significantly by COVID-19 acute infection severity. Individuals diagnosed with COVID-19 but never bedridden due to their illness were consistently at lower risks of depression and anxiety (PR 0·83, 95% CI 0·75-0·91 and 0·77, 0·63-0·94, respectively), while patients bedridden for more than 7 days were persistently at higher risks of symptoms of depression and anxiety (PR 1·61, 95% CI 1·27-2·05 and 1·43, 1·26-1·63, respectively) throughout the 16-month study period.

**CONCLUSION:** Acute infection severity is a key determinant of long-term mental morbidity among COVID-19 patients.

## Introduction

Adverse mental health symptoms^1-3^ and comorbid psychiatric disorders^3,4^ among coronavirus disease (COVID-19) patients have been extensively studied throughout the world. Meta-analyses suggest high symptom levels of depression and anxiety among COVID-19 patients, mostly based on patients within 6 months after hospital discharge^3,5,6^ In addition to the reported methodological shortcomings of the existing literature on this topic (i.e., inpatient samples and lack of comparison groups),^5,7^ little is still known about the long-term mental health of non-hospitalized COVID-19 patients with varying illness severities.

Both psychological and pathophysiological factors may contribute to mental health outcomes among patients diagnosed with COVID-19. First, the somewhat unpredictable disease course and prognosis,^8^ and worries of having infected others,^9^ along with extensive media coverage of traumatic effects of the disease^10^ may contribute to a transient rise in mental health symptoms. Second, the exacerbation of the COVID-19 illness resulting in severe flu-like symptoms and associated pro-inflammatory processes^4^ may contribute to the development of mental health symptoms among patients suffering the most severe disease course,^4,11^ for, as yet, unknown duration. Finally, several vulnerability factors, such as previous history of psychiatric disorders, may both be associated with the risk of a severe disease course after severe acute respiratory syndrome coronavirus 2 (SARS-CoV-2) infection^12^ and subsequent risks of adverse mental health symptoms.

Long or chronic COVID has been referred to as persistence of physical and mental symptoms beyond two months of infection.^13,14^ Yet, few studies have explored mental health symptom development beyond 6 months after diagnosis of COVID-19 in the general population as well as to what extent varying acute infection severity in COVID-19 predicts long-term mental health symptomology. To this end, we leveraged the multinational and population-based COVIDMENT cohorts^15^ to explore the mental health symptom trajectories up to 16 months after COVID-19 diagnosis. We hypothesized that mental health symptoms would decline with time from diagnosis while increased COVID-19 acute infection severity would be associated with long-term adverse mental health symptoms.

## Methods

### Study population and design

Seven cohorts from six countries were included in the present study (see Unnarsdóttir et al.^15^ for a comprehensive description) with harmonized data collections planned in April 2020. These were: The Danish Blood Donor Study (DBDS, N=71 562),^16^ The Estonian Biobank Covid-19 (EstBB-C19, N=14 452),^17^ The Icelandic COVID-19 National Resilience Cohort (C-19 Resilience, baseline, N=23 962),^18^ The Norwegian COVID-19, Mental Health and Adherence Project (MAP-19, N=10 061), The Norwegian Mother, Father and Child Cohort Study (MoBa, N=132 486),^19^ The Swedish Omtanke2020 (N=28 293), and The UK-based CovidLife (baseline, N=18 518).^20^ All cohorts had ethical approvals from respective national or regional ethics committees (see Supplemental Table S1) with varying numbers of waves of data collections from March 2020 through August 2021.

A total of 299 334 participants were eligible for analysis in all cohorts. After exclusion of participants with incomplete information on diagnosis of COVID-19 and main outcome measures of mental health symptoms, disease severity, and covariates, the analytic cohort consisted of 247 249 individuals (see Supplemental Figure S1 for a detailed flowchart for each cohort). We performed a cross-sectional analysis contrasting symptoms among individuals with and without a diagnosis of COVID-19 by time from diagnosis at the time of data collection. In a subpopulation with repeated symptom measures, we also tested potential change in symptom burden over time, from before and until after COVID-19 diagnosis.

### Measures

We used self-reports of a confirmed positive PCR-test (all cohorts) or positive antibody test (only in Omtanke2020) for SARS-CoV-2 infection as an indicator of a COVID-19 diagnosis (see Supplemental Table S1). Based on reported calendar month of diagnosis, the time from diagnosis until data collection was coded as 0—2 months (0—56 days); 2—6 months (57— 180 days); or 6—16 months (>180 days). To determine COVID-19 acute infection severity, we used the self-reports of the number of days bedridden due to COVID-19. In EstBB-C19 only, we used self-reports of the number of days with fever. We also obtained information on hospitalization for COVID-19 (see Supplemental Table S1 for further detail).

Several validated mental health instruments, including screening measures for depressive symptoms (Patient Health Questionnaire (PHQ-9) with the recommended cut-off of ≥10^21^ and Emotional State Questionnaire (EST-Q2) with the recommended cut-off of >11),^22,23^ anxiety (Angst-Symptom-Spørgeskemaet (ASS), EST-Q2^23^ and General Anxiety Disorder (GAD-7) with the recommended cut-off of ≥10),^24^ COVID-19 related distress (The Primary Care PTSD Screen for DSM-5 (PC-PTSD-5) with the recommended cut-off of ≥4,^25^ and the PTSD checklist for DSM-5 (PCL-5)),^26^ and sleep quality (EST-Q2,^23^ Pittsburgh Sleep Quality Index (PSQI))^27^ were included in all cohorts (see Supplemental Table S1 for further information). All the screening measures for posttraumatic stress were modified to refer specifically to COVID-19 (e.g., *‘Had nightmares about COVID-19?*’) and we therefore referred to this measure as COVID-19 related distress.

Covariates were the following (see Supplemental Table S1): Gender (male; female; other), age (continuous, in years), education (compulsory or less (no formal education); upper secondary, vocational, or other; bachelor’s/diploma university degree; master’s or Ph.D., not available in Omtanke2020), relationship status (in a relationship; single, not available in EstBB-C19), body mass index (BMI, <25, normal weight or underweight; 25—30, overweight; >30, obese), smoking status (never, former, current), history of diagnosis of any psychiatric disorder (yes; no), chronic medical condition (defined as hypertension, diabetes, heart disease, lung disease, chronic kidney disease (not available in DBDS), cancer, and immunosuppressive state (not available in DBDS) or immunosuppressive therapy (not available in DBDS) (no conditions; one condition; two conditions; > two conditions), response period (April June 2020; July September 2020; October December 2020; January March 2021; April August 2021).

### Statistical analysis

First, we explored the distribution of sociodemographic and health-related factors between individuals with and without a COVID-19 diagnosis for each cohort, individually and combined, along with symptom severity for individuals with a COVID-19 diagnosis. We then performed a cross-sectional analysis contrasting the prevalence of mental health indicators among individuals with and without a diagnosis of COVID-19, overall and in subgroup analyses by illness severity and time from diagnosis. Baseline measurements were used for individuals without a diagnosis of COVID-19 while the first measurement after diagnosis was used for the COVID-19 patients. We used robust (modified) Poisson regression models to estimate prevalence ratios (PRs) with 95% confidence intervals (95% CIs). We used classical sandwich estimator to control for intra-individual correlation when repeated measures were available.^28^ We adjusted the estimates for age and gender in the first model, and in the second model additionally for education, relationship status, smoking, BMI, previous psychiatric diagnosis, number of chronic medical conditions, and response period. Cohorts that did not measure specific covariates excluded them from their models (see Supplemental Table S1 for an overview).

Among the Icelandic C-19 Resilience, Norwegian MoBa, Swedish Omtanke2020, and the UK-based CovidLife cohorts, we performed a pair-wise comparison of repeated measures of mental health outcomes at two time-points among individuals with a COVID-19 diagnosis. Baseline measurements were the first response to a questionnaire after reporting a positive COVID-19 diagnosis, and the follow-up measurement was the last available response to a questionnaire. A generalised estimating equation (GEE) analysis was performed to test the difference of the mental health outcomes between the two time-points, while adjusting for response period. In addition, among individuals who answered at least one questionnaire before being diagnosed with COVID-19 and at least one questionnaire after being diagnosed, we performed a similar comparison of mental health measures at two time-points. The baseline measurement was the last questionnaire before a COVID-19 diagnosis and the follow-up measurement was the first questionnaire where a COVID-19 diagnosis was reported. A GEE analysis was performed to test the differences of the mental health outcomes between the two time-points, adjusting for response period.

We conducted meta-analyses based on aggregated data from each cohort with a random-effects model using the metafor package in R to estimate overall prevalence ratios for all the aforementioned analyses.^29^ Heterogeneity for each overall mental health outcome was examined using the I^2^-statistic.^30^ Statistical analyses were conducted in R (V.4.1.1), SAS 9.4, and Stata 17.0.

### Role of the funding source

The funder of the study had no role in study design, data collection, data analysis, data interpretation, or writing of the report.

## Results

### Background characteristics

Of the 247 249 participants in this study, 9979 (4%) reported having been diagnosed with COVID-19 at some point during the study period (Table 1). The proportion of COVID-19 varied across cohorts (see Supplemental Table S2) as did the cumulative prevalence of COVID-19 in each country during the study period (see Supplemental Table S3). A higher proportion of individuals with COVID-19 were female compared to those not diagnosed (67·9% vs. 61·7%). Individuals diagnosed with COVID-19 were younger than others (mean age: 46·6 vs. 48·9 years; Table 1). The mean age of the included participants in each cohort ranged from mid-40s to mid-50s, with the exception of the Norwegian MAP-19 cohort where mean age of participants was lower (33·3 years for COVID-19 patients vs. 35·7 years for others). Individuals diagnosed with COVID-19 had a higher educational level, were more likely to be in a relationship, have lower BMI, have a history of ever smoking, and chronic medical conditions, compared with individuals without a COVID-19 diagnosis (see Supplemental Table S2). Although the overall prevalence of history of psychiatric disorders varied considerably between individuals with and without a COVID-19 diagnosis (31·2% vs. 21·7%; Table 1), these differences were not considerable within each cohort (see Supplementary Table S2). Indeed, cohorts with a high proportion of individuals with COVID-19 (e.g., EstBB-C19 and Omtanke2020) also had a high percentage of previous diagnosis of psychiatric disorders both among individuals with and without a COVID-19 diagnosis.

**Table 1:**
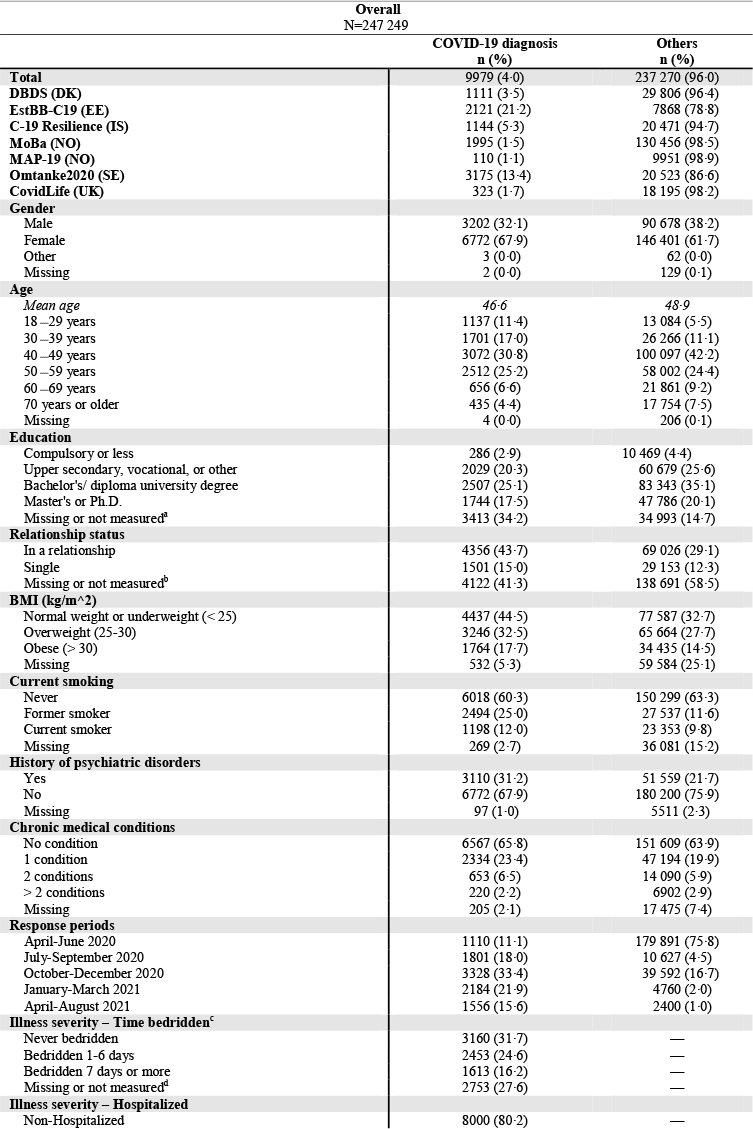
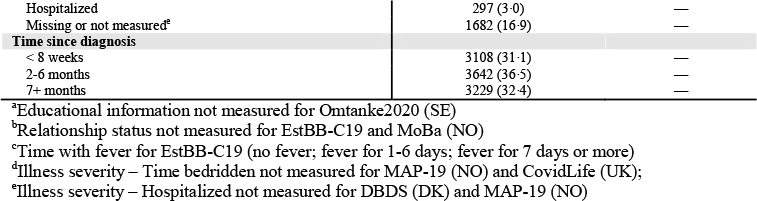
Background characteristics of study participants that received a COVID-19 diagnosis at any time point and others in the seven COVIDMENT cohorts.

### COVID-19 and mental health

Figure 1 (and Supplemental Table S4) shows prevalence ratios of adverse mental health symptoms among COVID-19 patients 0-16 months after diagnosis compared to individuals without such diagnosis, for each cohort and combined. The meta-analysis revealed that individuals with a COVID-19 diagnosis presented overall with a higher symptom burden of depression (PR 1·18, 95% CI 1·03-1·36; I^2^ 80.4%, p< 0·0001) and poor sleep quality (1·13, 1·03-1·24; I^2^ 76.8%, p=0·0028) but not anxiety (0·97, 0·91-1·03; I^2^ 0.0%, p=0·56) and COVID-19 related distress (1·05, 0·93-1·20; I^2^ 73.7%, p=0·0034; Figure 1). Estimates varied somewhat across all cohorts, especially for depression where results from EstBB-C19 differed significantly from other cohorts. When excluding EstBB-C19, the I^2^ measure of heterogeneity was no longer statistically significant (p=0·23). In the GEE analysis restricted to individuals who responded to at least one questionnaire before and one after being diagnosed with COVID-19 (median time between responses: C19-Resilience, 7·3 months; Omtanke2020, 1 month; CovidLife, 10 months), yielded similar prevalence ratios for depressive symptoms (PR 1·19, 95% CI 0·97-1·46; N=957) and poor sleep quality (1·09, 0·88-1·35; Supplemental Figure S2) after diagnosis of COVID-19, although with less precision.

**Figure 1:**
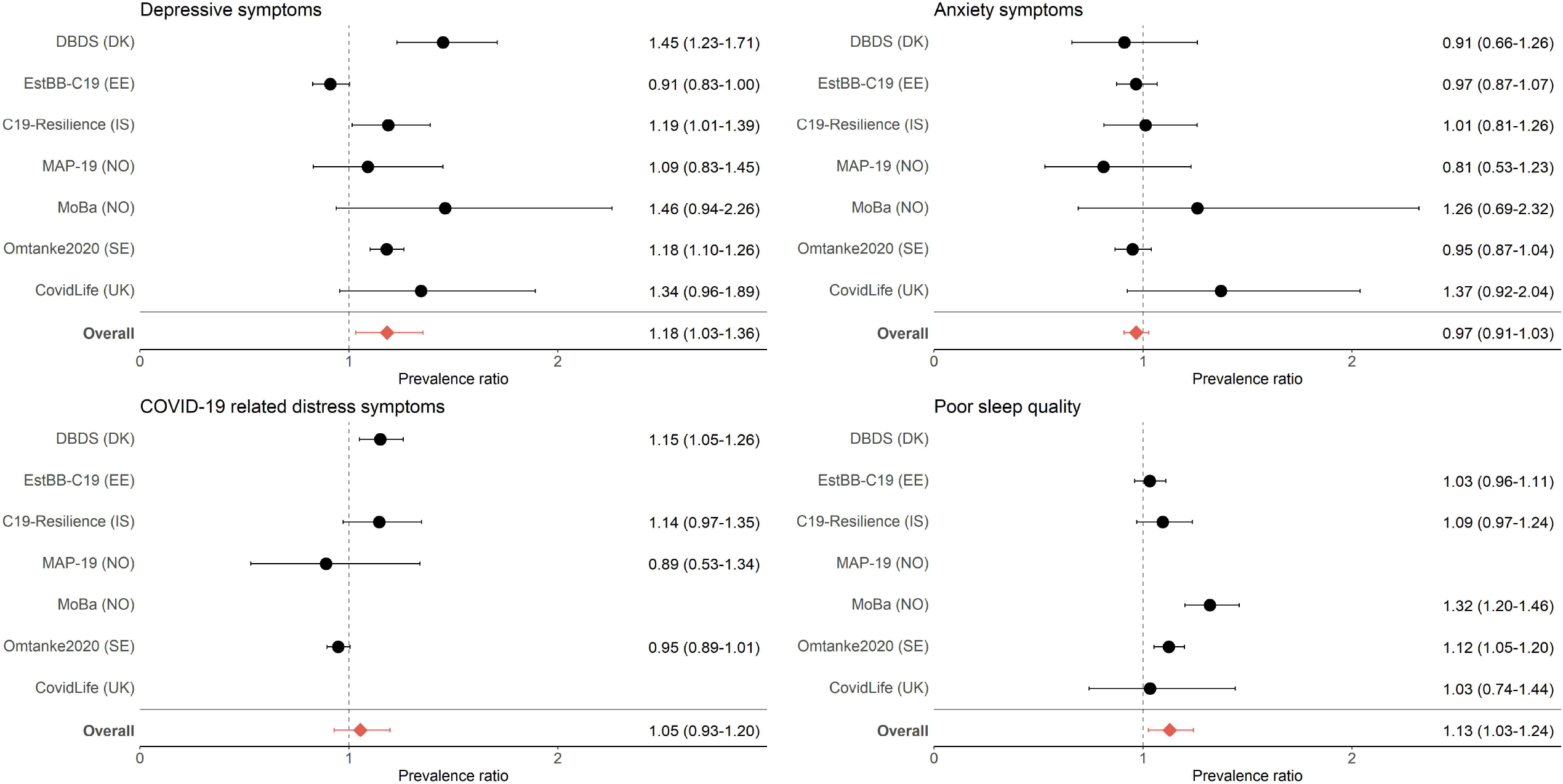
Prevalence ratios (with 95% confidence intervals) of mental health indicators among individuals with a diagnosis of COVID-19 compared with individuals without a COVID-19 diagnosis.

### Acute infection severity and mental health

Figure 2 (and Supplemental Table S4) shows prevalence ratios of mental morbidities among individuals diagnosed with COVID-19 between 0 and 16 months after diagnosis by time bedridden in their acute illness compared with those never diagnosed with COVID-19, for each cohort and combined. Longer time bedridden was consistently associated with increased prevalence ratios of all indicators of mental morbidities in a dose-response fashion. Compared with individuals never diagnosed with COVID-19, individuals who were never bedridden due to their SARS-CoV-2 infection had a significantly lower risk for symptoms of depression (PR 0·83, 95% CI 0·75-0·91) and anxiety (0·77, 0·63-0·94). Compared with individuals not diagnosed with COVID-19, individuals who were bedridden seven days and longer had a significantly higher risk of depressive symptoms (PR 1·61, 95% CI 1·27-2·05), anxiety symptoms (1·43, 1·26-1·63), and poor sleep quality (1·41, 1·24-1·61). In addition, a higher prevalence of COVID-19 related distress (PR 1·41, 95% CI 0·96-2·06) (Figure 2) was observed, although not statistically significant. Similarly, hospitalization (either non-ICU or ICU) was associated with increased prevalence ratios of all indicators of mental morbidities (Supplemental Figure S3). Overall, hospitalized COVID-19 patients who did not require ICU care were, compared with those never diagnosed with COVID-19, at a significantly increased risk for symptoms of depression (PR 1·80, 95% CI 1·47-2·20) and anxiety (1·47, 1·07-2·04), and poor sleep quality (1·49, 1·20-1·85). Patients admitted to the ICU were at increased risk for symptoms of anxiety (PR 1·57, 95% CI 1·05-2·34) and COVID-19 related distress (1·65, 1·06-2·58), when compared with individuals who were never diagnosed with COVID-19.

**Figure 2:**
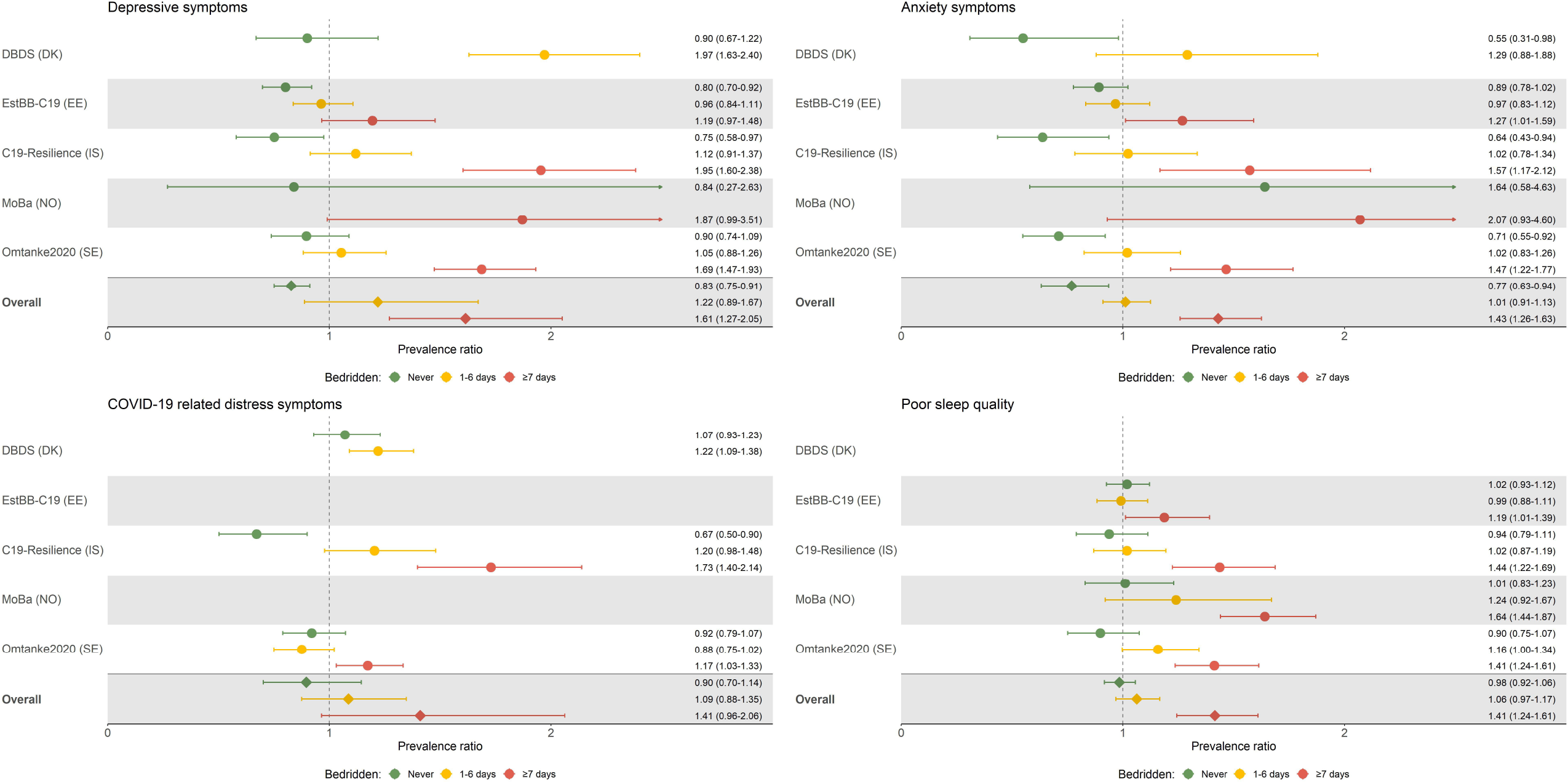
Mental health indicators among individuals with a diagnosis of COVID-19 compared with individuals without COVID-19 by illness severity (time bedridden).

### Mental health symptomology by time since diagnosis

Figure 3 (and Supplemental Table S4) shows the prevalence ratios of mental morbidities among individuals diagnosed with COVID-19 by time from diagnosis. Our meta-analysis suggests some attenuation of symptoms of depression and a clear attenuation of COVID-19 related distress with time from diagnosis. The prevalence ratio of depressive symptoms was 1·30 (95% CI 1·07-1·59) for individuals diagnosed within two months of assessment. The prevalence ratio was non-significant for individuals diagnosed with COVID-19 more than 2 months prior to the mental health assessment. For COVID-19 related distress, the prevalence ratio decreased from 1·20 (95% CI 1·09-1·33) within two months after diagnosis to non-significant levels beyond two months after diagnosis. These results were corroborated by estimates from the pair-wise comparison in the GEE analysis showing an attenuation in risks of symptoms of depression and poor sleep quality from first to last measurement after COVID-19 diagnosis (Supplemental Figure S4). Compared with the first measurement after diagnosis, the last measurement among COVID-19 patients, on average 9·7 months later, yielded a lower prevalence of depressive symptoms (PR 0·88, 95% CI 0·76-1·01; N=2883) and COVID-19 related distress (0·87, 0·75-1·00; N=3846). The time-dependent pattern for symptoms of anxiety and poor sleep quality was less clear (cf. Supplemental Figure S4).

**Figure 3:**
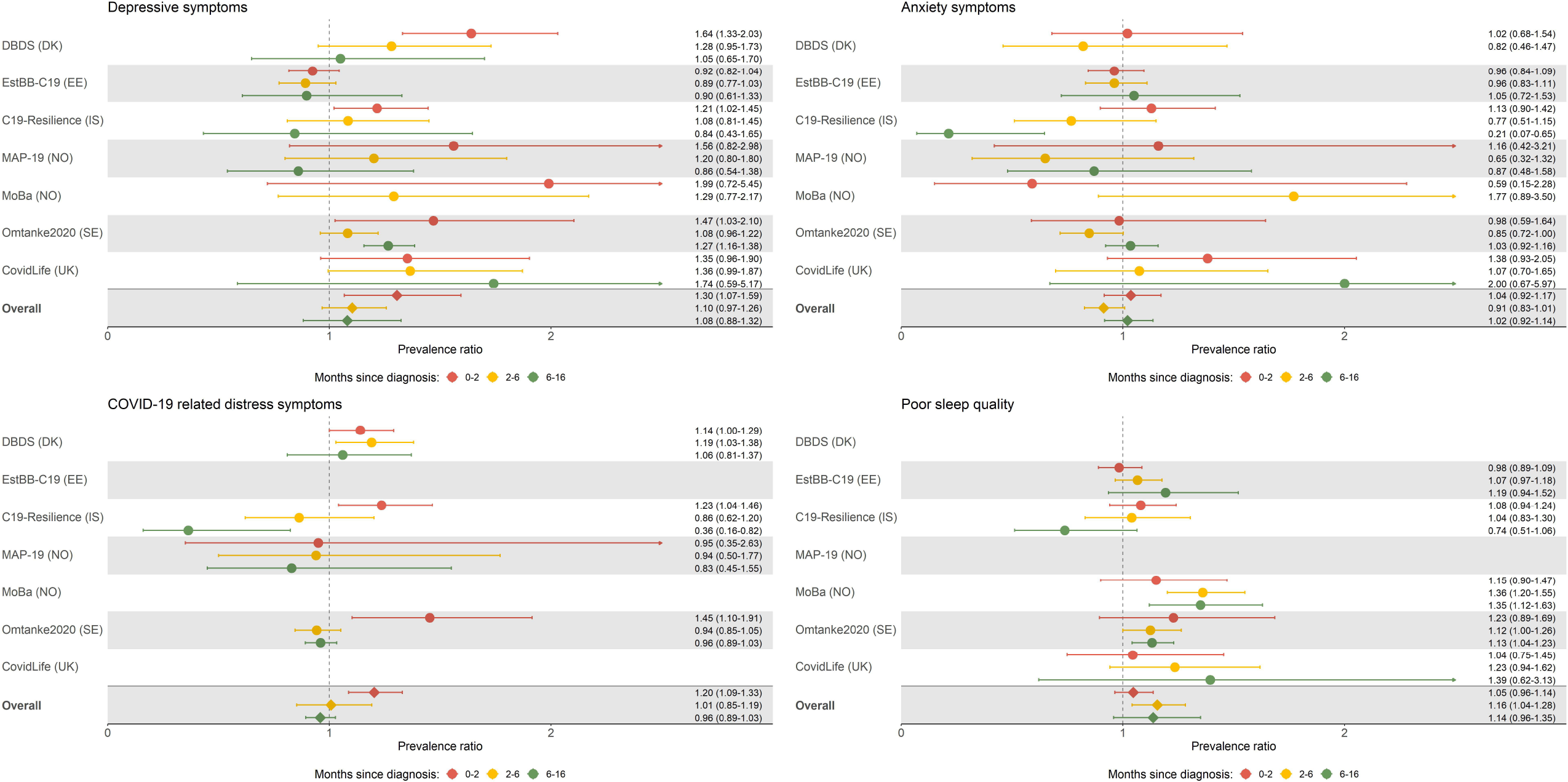
Mental health indicators among individuals with a diagnosis of COVID-19 compared with individuals without COVID-19 by time since diagnosis.

### Time trends by acute COVID-19 severity

Figure 4 shows 16-month mental health symptom development among individuals diagnosed with COVID-19 by time bedridden in the acute illness. While we found that mental health symptoms overall attenuated with time from COVID-19 diagnosis, patients bedridden for seven days or more showed persistent symptoms of depression (PRs 1·66, 95% CI 1·11-2·47; 1·53, 1·26-1·86; and 1·60, 1·17-2·81 at 0 2, 2 6, and 6 16 months post diagnosis, respectively) and anxiety (PRs 1·47, 95% CI 0·80-2·72; 1·46, 1·20-1·79; and 1·47, 1·19-1·81 at 0 2, 2 6, and 6 16 months post diagnosis, respectively), and poor sleep quality (PRs 1·35, 95% CI 1·11-1·64; 1·48, 1·29-1·69; and 1·26, 0·90-1·77 at 0 2, 2 6, and 6 16 months post diagnosis, respectively). In contrast, we observed a clear attenuation of COVID-19 related distress symptoms among COVID-19 patients bedridden for seven days or more (PRs 1·78, 95% CI 1·29-2·46; 1·17, 0·93-1·47; 1·01, 0·52-1·96 at 0 2, 2 6, and 6 16 months post diagnosis, respectively).

**Figure 4:**
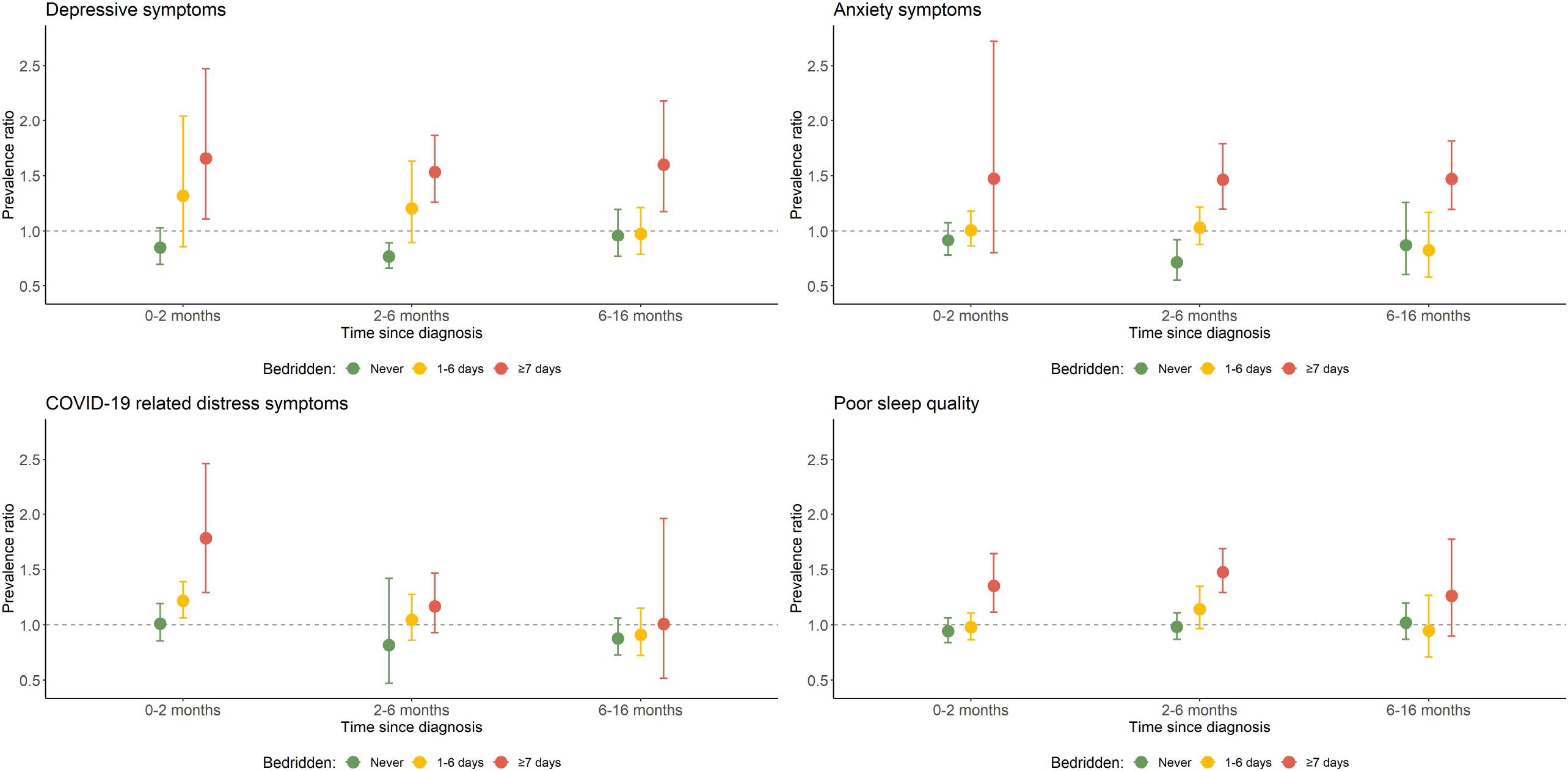
Mental health indicators during the first 16 months after diagnosis of COVID-19 by time bedridden.

## Discussion

In this multinational study including almost 250 000 individuals across six countries, we found that the acute COVID-19 severity is an important predictor of mental morbidities up to 16 months after diagnosis. While individuals who were never bedridden due to COVID-19 had lower risks of adverse mental health symptoms, extended time bedridden was associated with increased risk of mental morbidities in a dose-dependent manner. Thus, individuals bedridden due to COVID-19 illness for seven days or longer - representing 22% of the total patient population - persistently showed 50 60% increased risk of depression and anxiety symptoms throughout the first 16 months after diagnosis, while symptoms of COVID-19 related distress were significantly attenuated two months after diagnosis.

Since the start of the COVID-19 pandemic, our study is the first to report predictors of adverse mental health among COVID-19 infected individuals in the general population across six countries up to 16 months after infection. Our findings are in line with the limited existing literature, showing an adverse impact of COVID-19 diagnosis on mental-health status.^1-3,31^ However, the follow-up in these studies was at maximum six months, except for one recent study with one-year follow-up involving only hospitalized patients. In that study, 26% of COVID-19 patients had anxiety or depression at 12-month follow-up^31^ which is comparable to our study where 21% of those diagnosed with COVID-19 had depressive symptoms above cut-off at least six months after diagnosis (Supplemental Table S4). However, when limited to individuals hospitalized due to COVID-19 in our study, about 44% had symptoms of depression above cut-off 6-16 months post-diagnosis. In previous outbreaks of other infectious diseases, a high prevalence of posttraumatic stress disorder and depressive disorders have been observed up to 4 years after a diagnosis of severe acute respiratory syndrome (SARS) in 2003^32^ and these symptoms were also reported following the 2015 outbreak of Middle East Respiratory Syndrome (MERS) up to 12 months after diagnosis.^33^ Disease severity however was generally not addressed in these studies which also were limited to hospitalized patients.^33^

Our findings indicate that almost a quarter of the COVID-19 patient population had symptoms severe enough to be bedridden seven days or more and may specifically suffer persistent long-term mental health symptomology. The persistent elevated symptom levels of depression and anxiety among COVID-19 patients bedridden seven days or longer could be due to several mechanisms. This could include the worry of having infected others^9^ and the unpredictable prognosis of COVID-19 (e.g. worrying about long-term health effects, or even death^8^). However, such psychological mechanisms might be expected to lead to transient mental health symptoms that attenuate with time (i.e., when the patient and those that they may have infected recover from their illness). Indeed, the observed increase in symptoms of COVID-related distress in this group of patients may be indicative of such psychological mechanisms as the symptoms attenuated quickly after two months of diagnosis. In contrast, the persistent symptoms of depression and anxiety among individuals bedridden for seven days or longer may be due to continued physical long-COVID symptoms where functional limitation, loneliness, and lack of social contact (because of isolation) may cause worry and sense of helplessness.^13^ Alternatively, the inflammatory processes among patients suffering severe acute illness could also affect the risk of persistent mental health symptoms.^4,11^ Indeed, inflammation associated with chronic^34^ and infectious^35^ diseases have previously been linked with development of mental morbidities, particularly depression. Such mental morbidities have also been reported to persist after reduction in inflammation.^36^

While COVID-19 patients suffering a severe acute disease course had persistently increased risk of both anxiety and depression throughout the study period of the present study, individuals with a mild disease course had persistently lower risk of adverse mental health outcomes compared with the individuals without a COVID-19 diagnosis throughout the first year after infection. Several factors may contribute to this lower risk of mental morbidities among COVID-19 infected individuals with mild disease course. For example, individuals with a mild COVID-19 infection were able to return to somewhat more normal lives after the benign infection as compared with their more severely impacted counterparts who still could be restrained by fear of being infected in addition to official gathering restrictions. It is also possible that individuals with a low risk of mental morbidities before the pandemic, suffered less severe course after being infected with SARS-CoV-2, yielding the observed associations. Indeed, individuals with previous history of psychiatric disorders have been reported to be at increased risk of adverse COVID-19 outcomes.^37^ Although we did account for previous history of psychiatric disorders in the analysis, we cannot exclude the possibility of residual confounding.

The strengths of our study include the almost 10 000 individuals from the general population of six European countries who self-reported a confirmed diagnosis of COVID-19 of varying disease severities (ranging from virtually no COVID-19 symptoms to ICU admission) and the large comparison group without a COVID-19 diagnosis. The validated measures of mental health indicators along with the extended spectra of considered covariates diminish risks related to measurement bias and confounding. The limitations of our study include the self-reports of COVID-19 diagnosis and mental morbidities as well as the differences in response periods between individuals with and without COVID-19. About 75·8% of the comparison group responded between April and June 2020, while responses of patients with COVID-19 accumulated from April 2020 until August 2021. Yet, the response periods were included in the multivariable models and should therefore not explain our findings completely. Another possible weakness pertains to the different recruitment strategies of the included cohorts, which could explain the somewhat varying cohort specific results for some of the mental health outcomes. Some cohorts only recruited individuals from existing cohorts (e.g., MoBa, DBDS) and showed clearly lower overall symptom burden of mental morbidities than cohorts which used recruitment from both existing cohorts and open recruitment through social media (e.g., C19 Resilience, Omtanke2020, CovidLife). Moreover, 4% of the total multinational cohort reported a confirmed diagnosis of COVID-19, which is in line with the prevalence in four of the six countries included in this study (see Supplemental Table S3). Indeed, some national cohorts specifically targeted individuals diagnosed with COVID-19 in their recruitment (i.e., C19 Resilience and EstBB-C19) whilst others did not. Yet, on balance, the prevalence ratios were somewhat similar across cohorts, independent of recruitment strategies, indicating therefore absence of substantial systematic bias.

In conclusion, this multinational study identifies COVID-19 acute illness severity as a key determinant of long-term mental health symptomology among recovered patients. While COVID-19 patients with mild acute illness are unlikely to experience long-term mental health morbidities, more than a fifth of the included COVID-19 patients experienced severe acute illness course and subsequent persistent risk elevations of depressive and anxiety symptoms throughout 16 months after diagnosis. These findings motivate continued clinical surveillance and follow-up studies beyond the first year among individuals suffering the most severe symptomology after COVID-19 infections.

## Data Availability

The individual-level data underlying this article were subject to ethical approval and cannot be shared publicly due to data protection laws in each participating country.

## Data sharing

*The individual-level data underlying this article were subject to ethical approval and cannot be shared publicly due to* data protection laws in each participating country.

## Declaration of interests

This work was primarily supported with grants from Nordforsk (CovidMent, 105668) and Horizon2020 (CoMorMent, 847776). AM has received speakers’ fees from Illumina and Janssen and has received research grant funding from The Sackler Trust outside of the current work. OAA has received a grant to the institution from Nordforsk and Research Council of Norway for the present manuscript. OAA has also received a grant to the institution for an entity other than the present manuscript from South East Norway Health Authority, KG Jebsen Stiftelsen, and NIH, EU. OAA has received royalties for a textbook in psychiatry, consulting fees from HealthLytix and Milken Inst, payment for lectures from Sunovion and Lundbeck, and payment for expert testimony from the Norwegian Court. Outside of this work OAA has a patent for a devise for nasal delivery. OAA has participated at his institution on boards for local PI Clinical Trial Janssen, local PI Clinical Trial MAPS, and Local PI Clinical Trial Boehringer. UAV has received grants for the current work from Nordforsk and Horizon2020 as well as grants outside the current work from the Icelandic Research Fund, Swedish Research Council, Swedish Cancer Society and the European Research Council. All other authors declare no conflict of interest.

## Acknowledgements

This work was primarily supported with grants from Nordforsk (CovidMent, 105668) and Horizon2020 (CoMorMent, 847776).

EU H2020, CoMorMent, Nordforsk #CovidMent

Karina Banasik acknowledges the Novo Nordisk Foundation (grants NNF17OC0027594 and NNF14CC0001)

Helga Ask and Ragnar Nesvåg were supported by the Research Council of Norway (RCN 324 620), Per Magnus, Lill Trogstad and the MoBa Covid-19 data collection were supported by RCN (312 721), and Ole Andreassen was supported by RCN (273 291, 223 273).

Generation Scotland received core support from the Chief Scientist Office of the Scottish Government Health Directorates [CZD/16/6] and the Scottish Funding Council [HR03006] and is currently supported by the Wellcome Trust [216767/Z/19/Z]. Recruitment to the CovidLife study was facilitated by SHARE-the Scottish Health Research Register and Biobank. SHARE is supported by NHS Research Scotland, the Universities of Scotland and the Chief Scientist Office of the Scottish Government.

The research in the Estonian Biobank was supported by the European Union through the European Regional Development Fund (project no. 2014-2020.4.01.15-0012), and the Estonian Research Council through grant no. PSG615, the programme Mobilitas Pluss (MOBTP142), funding of Estonian sub-project of NordForsk project no. 105668, and National Programme for Addressing Socio-Economic Challenges through R&D (RITA), supported by the Estonian Government and European Regional Development Fund (RITA1/02-112).

## Author contributions

The COVIDMENT cohorts and/or their data collections were designed by IM, AL, ABU, DM, HA, KK, LANC, SUJ, AH, DJP, FF, JJ, KL, OAA, OBVP, TA, UAV and their respective teams. UAV, IM and TA directed the combined effort of this study implementation. UAV, IM, JJ and TA designed the analytical strategy in close collaboration with all team members and all authors helped to interpret the findings. IM, AL, ABU, DM, HA, KK, LANC, and SUJ conducted the literature review and drafted the manuscript under supervision of UAV. All authors revised the manuscript for critical content and approved the final version of the manuscript.

## Ethical approvals

The Danish Blood Donor Study (DBDS) was approved by the Zealand and Central Denmark Regional Committees on Health Research Ethics (Den Videnskabsetiske Komité for Region Sjælland and De Videnskabsetiske Komiteer for Region Midtjylland), Alléen 15, Sorø and Skottenborg 26, Viborg (SJ-740 and 1-10-72-95-13) and the Danish Data Protection Agency (Datatilsynet), Carl Jacobsens Vej 35, Valby (P-2019-99). The Estonian Biobank cohort (EstBB-C19) was approved by the Estonian Committee on Bioethics and Human Research (Eesti bioeetika ja inimuuringute nõukogu), Suur-Ameerika 1, 10122 Tallinn at Ministry of Social Affairs, Republic of Estonia (1.1-12/1277 and 1.1-12/2860). The Icelandic COVID-19 National Resilience Cohort (C-19 Resilience) was approved by the Icelandic National Bioethics Committee (Vísindasiðanefnd), Borgartún 21, Reykjavik (20-073 and 21-071) and Icelandic National Data Protection Authority (Persónuvernd), Rauðarárstíg 10, Reykjavik, Iceland. The Norwegian COVID-19, Mental Health and Adherence Project (MAP-19) was approved by the Regional Committee for Medical Research Ethics (Regionale komiteer for medisinsk og helsefaglig forskningsetikk), Gullhaugveien 1-3, Oslo in Norway (reference number: 125510) and Norwegian Centre for Research Data (Norsk senter for forskningsdata), Harald Hårfagres gate 29, Bergen, Norway (reference number: 802810). The Norwegian Mother, Father and Child Cohort Study (MoBa) was approved by The Regional Committee for Medical and Health Research Ethics (Regionale komiteer for medisinsk og helsefaglig forskningsetikk), Gullhaugveien 1-3, Oslo in Norway (127708/14140/20138). This also includes approval to link the MoBa data with data from national health registries (including psychiatric and COVID-19 diagnostic information). The Omtanke2020 study has been registered and approved by Swedish National Ethics Board (Etikprövningsmyndigheten), Box 2110, 750 02 Uppsala (DNR 2020-01785). The CovidLife study was reviewed and given a favourable opinion by the East of Scotland Research Ethics Committee in Dundee, Scotland (Reference: 20/ES/0021, AM02, AM04, AM05, AM11).

## Notes

### Competing Interest Statement

Competing interests: All authors have completed the ICMJE uniform disclosure form at www.icmje.org/coi_disclosure.pdf and declare:
This work was primarily supported with grants from Nordforsk (CovidMent, 105668) and Horizon2020 (CoMorMent, 847776). AM has received speakers' fees from Illumina and Janssen and has received research grant funding from The Sackler Trust outside of the current work. OAA has received a grant to the institution from Nordforsk and Research Council of Norway for the present manuscript. OAA has also received a grant to the institution for an entity other than the present manuscript from South East Norway Health Authority, KG Jebsen Stiftelsen, and NIH, EU. OAA has received royalties for a textbook in psychiatry, consulting fees from HealthLytix and Milken Inst, payment for lectures from Sunovion and Lundbeck, and payment for expert testimony from the Norwegian Court. Outside of this work OAA has a patent for a devise for nasal delivery. OAA has participated at his institution on boards for local PI Clinical Trial Janssen, local PI Clinical Trial MAPS, and Local PI Clinical Trial Boehringer. UAV has received grants for the current work from Nordforsk and Horizon2020 as well as grants outside the current work from the Icelandic Research Fund, Swedish Research Council, Swedish Cancer Society and the European Research Council. All other authors declare no conflict of interest.

### Author Declarations

The Danish Blood Donor Study (DBDS) was approved by the Zealand and Central Denmark Regional Committees on Health Research Ethics (Den Videnskabsetiske Komite for Region Sjaelland and De Videnskabsetiske Komiteer for Region Midtjylland), Alleen 15, Soro and Skottenborg 26, Viborg (SJ-740 and 1-10-72-95-13) and the Danish Data Protection Agency (Datatilsynet), Carl Jacobsens Vej 35, Valby (P-2019-99). The Estonian Biobank cohort (EstBB-C19) was approved by the Estonian Committee on Bioethics and Human Research (Eesti bioeetika ja inimuuringute noukogu), Suur-Ameerika 1, 10122 Tallinn at Ministry of Social Affairs, Republic of Estonia (1.1-12/1277 and 1.1-12/2860). The Icelandic COVID-19 National Resilience Cohort (C-19 Resilience) was approved by the Icelandic National Bioethics Committee (Visindasidanefnd), Borgartun 21, Reykjavik (20-073 and 21-071) and Icelandic National Data Protection Authority (Personuvernd), Raudararstig 10, Reykjavik, Iceland. The Norwegian COVID-19, Mental Health and Adherence Project (MAP-19) was approved by the Regional Committee for Medical Research Ethics (Regionale komiteer for medisinsk og helsefaglig forskningsetikk), Gullhaugveien 1-3, Oslo in Norway (reference number: 125510) and Norwegian Centre for Research Data (Norsk senter for forskningsdata), Harald Harfagres gate 29, Bergen, Norway (reference number: 802810). The Norwegian Mother, Father and Child Cohort Study (MoBa) was approved by The Regional Committee for Medical and Health Research Ethics (Regionale komiteer for medisinsk og helsefaglig forskningsetikk), Gullhaugveien 1-3, Oslo in Norway (127708/14140/20138). This also includes approval to link the MoBa data with data from national health registries (including psychiatric and COVID-19 diagnostic information). The Omtanke2020 study has been registered and approved by Swedish National Ethics Board (Etikprovningsmyndigheten), Box 2110, 750 02 Uppsala (DNR 2020-01785). The CovidLife study was reviewed and given a favourable opinion by the East of Scotland Research Ethics Committee in Dundee, Scotland (Reference: 20/ES/0021, AM02, AM04, AM05, AM11).

