## Supplemental material for "Acute COVID-19 severity and 16-month mental morbidity trajectories in patient populations of six nations"

### Supplementary Material

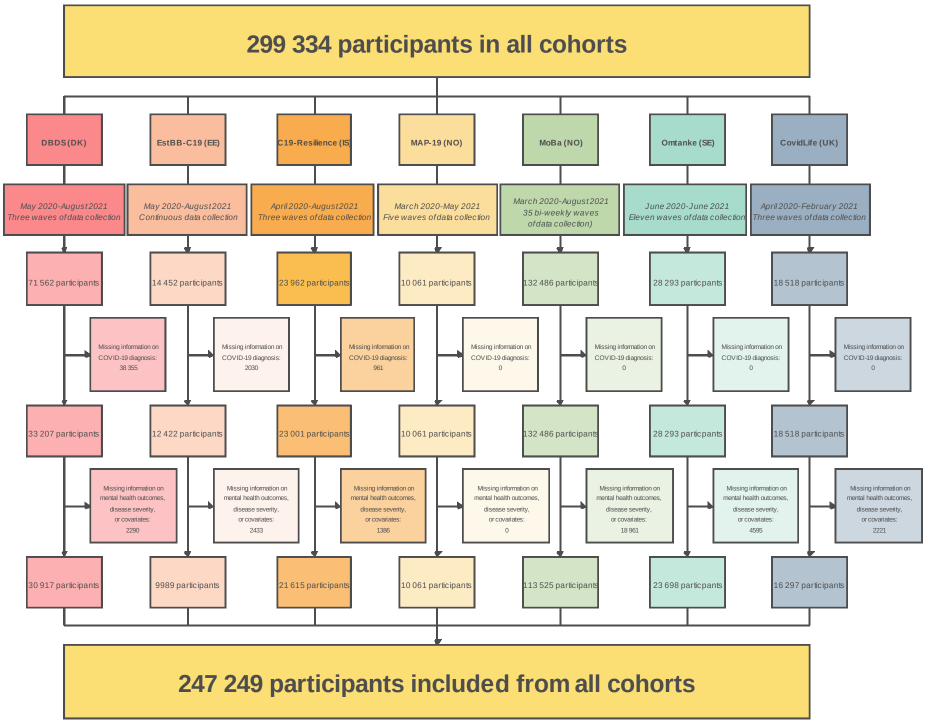

Figure S1: **Flowchart of participation and exclusions in each cohort yielding the study population.**

Table S1: **Overview of information and various measures used in study**

| | **Denmark** DBDS | **Estonia** EstBB-C19 | **Iceland** C19-Resilience | **Norway** MAP-19 | **Norway** MoBa | **Sweden** Omtanke2020 | **UK** CovidLife |
|---|---|---|---|---|---|---|---|
| | | | | **Ethical approvals** | | | |
| | Zealand and Central Denmark Regional Committees on Health Research Ethics (SJ-740 and 1-10-72-95-13) and the Data Protection Agency (P-2019–99) | Estonian Committee on Bioethics and Human Research (1.1–12/1277) | National Bioethics Committee (NBC no. 20–073, 21–071) as well as the National Data Protection Authority | Regional Committee for Medical Research Ethics, reference number: 125510 | Regional Committees for Medical and Health Research Ethics (127708/14140/20138) | Ethical approval no. 2020–01785 | East of Scotland Research Ethics Service (EoSRES) |
| | | | | **COVID-19 related outcomes** | | | |
| **COVID-19 diagnosis** | Self-report of confirmed PCR-test | Self-report of confirmed PCR-test | Self-report of confirmed PCR-test | Self-report of confirmed PCR-test | Self-report of confirmed PCR-test | Self-report of confirmed PCR-test or positive antibody test with date of testing provided | Self-report of confirmed PCR-test |
| **COVID-19 illness severity** | Never bedridden/Bedridden 1 day or more | No fever/ Fever 1-6 days/Fever 7 days or more | Never bedridden/Bedridden 1-6 days/Bedridden 7 days or more | Not measured | Never bedridden/Bedridden 1-6 days/Bedridden 7 days or more | Never bedridden/Bedridden 1-6 days/Bedridden 7 days or more (only asked at long surveys (baselin and 6-months FU) | Not measured |
| **COVID-19 hospitalization** | Not measured | Non-hospitalized/ Hospitalized (non-ICU)/Hospitalized (ICU) | Non-hospitalized/ Hospitalized (non-ICU)/Hospitalized (ICU) | Not measured | Non-hospitalized/ Hospitalized | Non-hospitalized/Hospitalized (non-ICU)/Hospitalized (ICU) | Non-hospitalized/ Hospitalized (non-ICU)/Hospitalized (ICU) |
| | | | | **Mental health instruments** | | | |
| **Depression** | PHQ-9 | EST-Q2 | PHQ-9 | PHQ-9 | PHQ-9 | PHQ-9 | PHQ-9 |
| **Anxiety** | ASS | EST-Q2 | GAD-7 | GAD-7 | GAD-7 | GAD-7 | GAD-7 |
| **COVID-19 related distress** | PC-PTSD-5 (modified) | Not measured | PC-PTSD-5 (modified) | PCL-5 | PC-PTSD-5 (modified) | PC-PTSD-5 (modified) | Not measured |
| **Sleep quality** | Not measured | EST-Q2 | PSQI | Not measured | PSQI | One item of PSQI | Binary variable of better/same sleep quality versus worse sleep quality |
| | | | | **Covariates** | | | |
| **Age** | x | x | x | x | x | x | x |
| **Gender** | x | x | x | x | x | x | x |
| **Education** | x | x | x | x | x | — | x |
| **Relationship status** | x | — | x | x | — | x | x |
| **Smoking** | x | x | x | x | x | x | x |
| **BMI** | x | x | x | x | x | x | x |
| **Pre-existing psychiatric conditions** | * | * | x | x | x | x | x |
| **Number of chronic medical conditions** | x | x | x | — | x | x | x |
| **Response period** | x | x | x | x | x | x | x |

Note: Data from surveys are marked with x; register data is marked with a *. “x” and “*” denotes covariates were included in model while “—” indicates covariate was not included

Table S2: **Background characteristics of study participants that received a COVID-19 diagnosis at any time point and others in the seven COVIDMENT cohorts separately, combined, and combined with all missing values removed**

| | Denmark | | Estonia | | Iceland | | Norway | | | | Sweden | | United Kingdom | | Overall | | Overall | |
|---|---|---|---|---|---|---|---|---|---|---|---|---|---|---|---|---|---|---|
| | (DBDS) | | (EstBB C-19) | | (C19-Resilience) | | (MAP-19) | | (MoBa) | | (Omtanke) | | (CovidLife) | | | | without missing values | |
| | (N=30 917) | | (N=9989) | | (N=21 615) | | (N = 10 061) | | (N=132 451) | | (N=23 698) | | (N=18 518) | | (N= 247 249) | | (N=247 249) | |
| | **COVID-19 diagnosis** | **Others** | **COVID-19 diagnosis** | **Others** | **COVID-19 diagnosis** | **Others** | **COVID-19 diagnosis** | **Others** | **COVID-19 diagnosis** | **Others** | **COVID-19 diagnosis** | **Others** | **COVID-19 diagnosis** | **Others** | **COVID-19 diagnosis** | **Others** | **COVID-19 diagnosis** | **Others** |
| | **n (%)** | **n (%)** | **n (%)** | **n (%)** | **n (%)** | **n (%)** | **n (%)** | **n (%)** | **n (%)** | **n (%)** | **n (%)** | **n (%)** | **n (%)** | **n (%)** | **n (%)** | **n (%)** | **n (%)** | **n (%)** |
| **Total** | 1111 (3.5) | 29 806 (96.4) | 2121 (21.2) | 7868 (78.8) | 1144 (5.3) | 20 471 (94.7) | 110 (1.1) | 9951 (98.9) | 1995 (1.5) | 130 456 (98.5) | 3175 (13.4) | 20 523 (86.6) | 323 (1.7) | 18 195 (98.2) | 9979 (4) | 237270 (96) | 9979 (4) | 237270 (96) |
| **Gender** | | | | | | | | | | | | | | | | | | |
| Male | 513 (46.2) | 14 341 (48.1) | 740 (34.9) | 2224 (28.3) | 424 (37.1) | 6125 (29.9) | 17 (15.5) | 2166 (21.8) | 815 (40.9) | 56 098 (43.0) | 607 (19.1) | 3796 (18.5) | 86 (26.6) | 5928 (32.6) | 3202 (32.1) | 90678 (38.2) | 3202 (32.1) | 90678 (38.2) |
| Female | 598 (53.8) | 15 465 (51.9) | 1381 (65.1) | 5664 (71.7) | 718 (62.8) | 14 309 (69.9) | 92 (83.6) | 7760 (78.0) | 1180 (59.1) | 74 358 (57.0) | 2568 (80.9) | 16 727 (81.5) | 235 (72.8) | 12 138 (66.7) | 6772 (67.9) | 146401 (61.7) | 6772 (67.9) | 146401 (61.7) |
| Other | — | — | — | — | 2 (0.2) | 37 (0.2) | 1 (0.9) | 25 (0.3) | — | — | ** | ** | — | — | 3 (0) | 62 (0) | 3 (0) | 62 (0) |
| Missing | — | — | — | — | — | — | — | — | — | — | — | — | 2 (0.6) | 129 (0.7) | 2 (0) | 129 (0.1) | | |
| **Age** | | | | | | | | | | | | | | | | | | |
| *Mean age (SD)* | *46.6 (14.8)* | *54.6 (16.6)* | *44.5 (13.4)* | *44.7 (13.3)* | *48.2 (14.7)* | *54.8 (14.1)* | *33.3 (12.2)* | *35.7 (13.5)* | *47.1 (5.41)* | *46.7 (5.39)* | *47.1 (13.8)* | *50.4 (16.1)* | *52.4 (13.5)* | *56.5 (14.3)* | *46.6* | *48.9* | *46.6* | *48.9* |
| 18-29 years | 200 (18.0) | 3053 (10.2) | 273 (12.9) | 970 (12.3) | 168 (14.7) | 1216 (5.9) | 54 (49.1) | 4349 (43.7) | — | — | 419 (13.2) | 2532 (12.3) | 23 (7.1) | 964 (5.3) | 1137 (11.4) | 13084 (5.5) | 1137 (11.4) | 13084 (5.5) |
| 30-39 years | 194 (17.5) | 3554 (11.9) | 573 (27.0) | 2143 (27.2) | 154 (13.5) | 1974 (9.6) | 23 (20.9) | 2254 (22.7) | 154 (7.7) | 11 039 (8.5) | 565 (17.8) | 3542 (17.3) | 38 (11.8) | 1760 (9.7) | 1701 (17) | 26266 (11.1) | 1701 (17.1) | 26266 (11.1) |
| 40-49 years | 261 (23.5) | 4911 (16.5) | 563 (26.5) | 1963 (24.9) | 259 (22.6) | 3620 (17.7) | 21 (19.1) | 1567 (15.7) | 1232 (61.8) | 81 932 (62.8) | 677 (21.3) | 3596 (17.5) | 59 (18.3) | 2508 (13.8) | 3072 (30.8) | 100097 (42.2) | 3072 (30.8) | 100097 (42.2) |
| 50-59 years | 244 (22.0) | 6152 (20.5) | 396 (18.7) | 1607 (20.4) | 284 (24.8) | 5280 (25.8) | 6 (5.5) | 1088 (10.9) | 575 (28.8) | 35 620 (27.3) | 908 (28.6) | 4300 (21.0) | 99 (30.7) | 3955 (21.7) | 2512 (25.2) | 58002 (24.4) | 2512 (25.2) | 58002 (24.5) |
| 60-69 years | 117 (10.5) | 4386 (14.7) | 221 (10.4) | 856 (10.9) | 206 (18.0) | 5335 (26.1) | 5 (4.5) | 557 (5.6) | 33 (1.7) | 1768 (1.4) | 462 (14.6) | 3505 (17.1) | 74 (22.9) | 5454 (30.0) | 1118 (11.2) | 21861 (9.2) | 1118 (11.2) | 21861 (9.2) |
| 70 years + | 95 (8.6) | 7750 (26.0) | 95 (4.5) | 329 (4.2) | 73 (6.4) | 3046 (14.9) | 1 (0.9) | 136 (1.4) | 1 (0.1) | 97 (0.1) | 144 (4.5) | 3048 (14.9) | 26 (8.0) | 3348 (18.4) | 435 (4.4) | 17754 (7.5) | 435 (4.4) | 17754 (7.5) |
| Missing | | | | | | | | | | | | | 4 (1.2) | 206 (1.1) | 4 (0) | 206 (0.1) | | |
| **Education** | | | | | | | | | | | | | | | | | | |
| Compulsory | 30 (2.7) | 1501 (5.0) | 44 (2.1) | 140 (1.8) | 144 (12.6) | 2916 (14.2) | 3 (2.7) | 519 (5.2) | 38 (1.9) | 3498 (2.7) | * | * | 23 (7.1) | 1515 (8.3) | 286 (2.9) | 10469 (4.4) | 290 (4.4) | 10469 (5.2) |
| Upper secondary, vocational or other | 60 (5.4) | 1144 (3.8) | 884 (41.7) | 2825 (35.9) | 361 (31.6) | 6359 (31.1) | 15 (13.6) | 1771 (17.8) | 606 (30.4) | 42 530 (32.6) | * | * | 103 (31.9) | 6050 (33.3) | 2029 (20.3) | 60679 (25.6) | 2029 (30.9) | 60679 (30) |
| Bachelor's/ diploma university degree | 698 (62.8) | 20 142 (67.6) | 582 (27.4) | 2182 (27.7) | 390 (34.1) | 6473 (31.6) | 92 (83.6)*1 | 7661 (77.0) *1 | 661 (33.1) | 42 765 (32.8) | * | * | 84 (26.0) | 4120 (22.6) | 2507 (25.1) | 83343 (35.1) | 2507 (38.2) | 83343 (41.2) |

| | | | | | | | | | | | | | | | | | | |
|---|---|---|---|---|---|---|---|---|---|---|---|---|---|---|---|---|---|---|
| Master's or Ph.D. | 323 (29.1) | 7019 (23.5) | 611 (28.8) | 2721 (34.6) | 249 (21.8) | 4723 (23.1) | | | 479 (24.0) | 28 879 (22.1) | * | * | 82 (25.4) | 4444 (24.4) | 1744 (17.5) | 47786 (20.1) | 1744 (26.5) | 47786 (23.6) |
| No formal education | — | — | — | — | — | — | | | — | — | * | * | 4 (1.2) | 380 (2.1) | 3413 (34.2) | 34993 (14.7) | | |
| Missing | — | — | — | — | — | — | | | 211 (10.6) | 12 784 (9.8) | * | * | 27 (8.4) | 1686 (9.3) | | | | |
| **Marital status** | | | | | | | | | | | | | | | | | | |
| In a relationship | 735 (66.2) | 18 901 (63.4) | — | — | 910 (79.5) | 15 779 (77.1) | 64 (58.2) | 5590 (60.2) | — | — | 2409 (75.9) | 15 034 (73.3) | 238 (73.7) | 13 722 (75.4) | 4356 (43.7) | 69026 (29.1) | 4356 (74.4) | 69026 (70.3) |
| Single | 376 (33.8) | 10 905 (36.6) | — | — | 234 (20.5) | 4692 (22.9) | 46 (41.8) | 3961 (39.8) | — | — | 766 (24.1) | 5489 (26.7) | 79 (24.5) | 4106 (22.6) | 1501 (15) | 29153 (12.3) | 1501 (25.6) | 29153 (29.7) |
| Missing | — | — | — | — | — | — | | | — | — | — | — | 6 (1.9) | 367 (2.0) | 4122 (41.3) | 138691 (58.5) | | |
| **BMI (kg/m^2)** | | | | | | | | | | | | | | | | | | |
| < 25, Normal weight | 548 (49.3) | 13 441 (45.1) | 980 (46.2) | 3710 (47.2) | 378 (33.0) | 6028 (29.4) | 33 (30.0) | 1801 (18.1) | 651 (32.6) | 34 256 (26.3) | 1733 (75.9) | 11 458 (55.8) | 114 (35.3) | 6893 (37.9) | 4437 (44.5) | 77587 (32.7) | 4437 (47) | 77587 (43.7) |
| 25-30, Overweight | 414 (37.3) | 11 461 (38.5) | 704 (33.2) | 2521 (32.0) | 434 (37.9) | 8110 (39.6) | 22 (20.0) | 1405 (14.1) | 570 (28.6) | 29 725 (22.8) | 992 (31.2) | 6341 (30.9) | 110 (34.1) | 6101 (33.5) | 3246 (32.5) | 65664 (27.7) | 3246 (34.4) | 65664 (37) |
| > 30, Obese | 149 (13.4) | 4904 (16.5) | 437 (20.6) | 1637 (20.8) | 332 (29.0) | 6333 (30.9) | 11 (10.0) | 695 (7.0) | 298 (14.9) | 13 575 (10.4) | 450 (14.2) | 2724 (13.3) | 87 (26.9) | 4567 (25.1) | 1764 (17.7) | 34435 (14.5) | 1764 (18.7) | 34435 (19.4) |
| Missing | — | — | — | — | — | — | 44 (40.0) | 6050 (60.8) | 476 (23.9) | 52 900 (40.6) | — | — | 12 (3.7) | 634 (3.5) | 532 (5.3) | 59584 (25.1) | | |
| **Current smoking** | | | | | | | | | | | | | | | | | | |
| No, never | 840 (75.6) | 21 662 (72.7) | 1130 (53.3) | 4210 (53.5) | 565 (49.4) | 9441 (46.1) | 47 (42.7) | 2934 (29.5) | 1643 (82.4) | 90 547 (69.4) | 1606 (50.6) | 10 748 (52.4) | 187 (57.9) | 10 757 (59.1) | 6018 (60.3) | 150299 (63.3) | 6018 (62) | 150299 (74.7) |
| No, former smoker | 198 (17.8) | 5252 (17.6) | 722 (34.0) | 2515 (32.0) | 421 (36.8) | 7989 (39.0) | | | - | - | 1051 (33.1) | 6415 (31.3) | 102 (31.6) | 5366 (29.5) | 2494 (25) | 27537 (11.6) | 2494 (25.7) | 27537 (13.7) |
| Yes, currently | 73 (6.6) | 2892 (9.7) | 269 (12.7) | 1143 (14.5) | 158 (13.8) | 3041 (14.9) | 32 (29.1) | 1922 (19.3) | 129 (6.5) | 9697 (7.4) | 518 (16.3) | 3360 (16.4) | 19 (5.9) | 1298 (7.1) | 1198 (12) | 23353 (9.8) | 1198 (12.3) | 23353 (11.6) |
| Missing | — | — | — | — | — | — | 31 (28.2) | 5095 (51.2) | 223 (11.2) | 30 212 (23.2) | — | — | 15 (4.6) | 774 (4.3) | 269 (2.7) | 36081 (15.2) | | |
| **History of psychiatric disorders** | | | | | | | | | | | | | | | | | | |
| Yes | 211 (19.0)* | 7059 (23.7)* | 1094 (51.6) | 3789 (48.2) | 287 (25.1) | 5927 (29.0) | 17 (15.5) | 1702 (17.1)*2 | 336 (16.8) | 20 465 (15.7) | 1056 (33.3) | 6700 (32.6) | 109 (33.7) | 5917 (32.5) | 3110 (31.2) | 51559 (21.7) | 3110 (31.5) | 51559 (22.2) |
| No | 900 (81.0)* | 22 747 (76.3)* | 1027 (48.4) | 4079 (51.8) | 857 (74.9) | 14 544 (71.0) | 93 (84.5) | 8249 (82.9) | 1581 (79.2) | 105 084 (80.6) | 2104 (66.3) | 13 618 (66.4) | 210 (650) | 11 879 (65.3) | 6772 (67.9) | 180200 (75.9) | 6772 (68.5) | 180200 (77.8) |
| Missing | — | — | — | — | — | — | | | 78 (3.9) | 4907 (3.8) | 15 (0.5) | 205 (1.0) | 4 (1.2) | 399 (2.2) | 97 (1) | 5511 (2.3) | | |
| **Chronic medical conditions** | | | | | | | | | | | | | | | | | | |
| No condition | 500 (45.0) | 10 151 (34.1) | 1284 (60.5) | 4359 (55.4) | 785 (68.6) | 12 021 (58.7) | - | - | 1541 (77.2) | 101 521 (77.8) | 2288 (72.1) | 13 899 (67.7) | 169 (52.3) | 9658 (53.1) | 6567 (65.8) | 151609 (63.9) | 6567 (67.2) | 151609 (69) |
| 1 condition | 357 (32.1) | 9652 (32.4) | 604 (28.5) | 2565 (32.6) | 274 (24.0) | 5968 (29.2) | - | - | 311 (15.6) | 19 014 (14.6) | 684 (21.5) | 4867 (23.7) | 104 (32.2) | 5128 (28.2) | 2334 (23.4) | 47194 (19.9) | 2334 (23.9) | 47194 (21.5) |
| 2 conditions | 188 (16.9) | 5820 (19.5) | 171 (8.1) | 653 (8.3) | 60 (5.2) | 1908 (9.3) | - | - | 43 (2.2) | 2364 (1.8) | 161 (5.1) | 1303 (6.3) | 30 (9.3) | 2042 (11.2) | 653 (6.5) | 14090 (5.9) | 653 (6.7) | 14090 (6.4) |
| > 2 conditions | 66 (5.9) | 4183 (14.0) | 62.0 (2.9) | 291 (3.7) | 25 (2.2) | 574 (2.8) | - | - | 9 (0.5) | 360 (0.3) | 42 (1.3) | 454 (2.2) | 16 (5.0) | 1040 (5.7) | 220 (2.2) | 6902 (2.9) | 220 (2.3) | 6902 (3.1) |
| Missing | — | — | — | — | — | — | - | - | 91 (4.6) | 7197 (5.5) | — | — | 4 (1.2) | 327 (1.8) | 205 (2.1) | 17475 (7.4) | | |
| **Response periods** | | | | | | | | | | | | | | | | | | |

| | | | | | | | | | | | | | | | | | | |
|---|---|---|---|---|---|---|---|---|---|---|---|---|---|---|---|---|---|---|
| April-June 2020 | 125 (11.3) | 52 (0.2) | 138 (6.5) | 1113 (14.1) | 372 (32.5) | 19 807 (96.8) | 14 (12.7)*3 | 9951 (100) | 346 (17.3) | 130 456 (100) | 45 (1.4) | 317 (1.5) | 70 (21.7) | 18 195 (100) | 1110 (11.1) | 179891 (75.8) | 1110 (11.1) | 179891 (75.8) |
| July-September 2020 | — | — | 155 (7.3) | 1133 (14.4) | 9 (0.8) | 232 (1.1) | 3 (2.7) | - | 292 (14.6) | - | 1322 (41.6) | 9262 (45.1) | 20 (6.2) | — | 1801 (18) | 10627 (4.5) | 1801 (18) | 10627 (4.5) |
| October-December 2020 | 947 (85.2) | 29 049 (97.5) | 166 (7.8) | 1328 (16.9) | 139 (12.2) | 396 (1.9) | 26 (23.6) | - | 682 (34.2) | - | 1368 (43.1) | 8819 (43.0) | — | — | 3328 (33.4) | 39592 (16.7) | 3328 (33.4) | 39592 (16.7) |
| January-March 2021 | 39 (3.5) | 705 (2.4) | 905 (42.7) | 2499 (31.8) | 574 (50.2) | 32 (0.2) | 28 (25.5) | - | 86 (4.3) | - | 319 (10.0) | 1524 (7.4) | 233 (72.1) | — | 2184 (21.9) | 4760 (2) | 2184 (21.9) | 4760 (2) |
| April-June 2021 | — | — | 757 (35.7) | 1795 (22.8) | 50 (4.4) | 4 (0.0) | 39 (35.5) | - | 589 (29.5) | - | 121 (3.8) | 601 (2.9) | — | — | 1556 (15.6) | 2400 (1) | 1556 (15.6) | 2400 (1) |
| **Illness severity** | | | | | | | | | | | | | | | | | | |
| Never bedridden | 560 (50.4) | — | 1061 (50.0) | — | 428 (37.4) | — | - | - | 636 (31.9) | — | 475 (15.0) | — | — | — | 3160 (31.7) | — | 3160 (43.7) | — |
| Bedridden 1-6 days | 551 (49.6) | — | 773 (36.4) | — | 455 (39.8) | — | - | - | 213 (10.7) | — | 461 (14.5) | — | — | — | 2453 (24.6) | — | 2453 (33.9) | — |
| Bedridden 7 days or more | – | — | 287 (13.5) | — | 261 (22.8) | — | - | - | 685 (34.3) | — | 380 (12.0) | — | — | — | 1613 (16.2) | — | 1613 (22.3) | — |
| Missing | — | — | — | — | — | — | - | - | 461 (23.1) | — | 1859 (58.6) | — | — | — | 2753 (27.6) | — | | — |
| Non-Hospitalized | – | — | 2073 (97.7) | — | 1056 (92.3) | — | - | - | 1495 (74.9) | — | 3090 (97.3) | — | 286 (88.5) | — | 8000 (80.2) | — | 8000 (96.4) | |
| Hospitalized | – | — | 48 (2.3) | — | 88 (7.7) | — | - | - | 39 (2.0) | — | 85 (0.3) | — | 37 (11.4) | — | 297 (3) | — | 297 (3.6) | — |
| Missing | — | — | — | — | — | — | - | - | 461 (23.1) | — | — | — | — | — | 1682 (16.9) | — | | |
| **Time since diagnosis** | | | | | | | | | (at wave 28) | | | | | | | | | — |
| 0-2 months | 594 (53.5) | — | 1055 (49.7) | — | 444 (38.8) | — | 17 (15.5) | - | 568 (28.5) | — | 215 (6.8) | — | 215 (66.6) | — | 3108 (31.1) | — | 3108 (31.1) | — |
| 2-6 months | 373 (33.6) | — | 954 (45.0) | — | 541 (47.3) | — | 43 (39.1) | - | 951 (47.7) | — | 682 (21.5) | — | 98 (30.3) | — | 3642 (36.5) | — | 3642 (36.5) | — |
| 6-16 months | 144 (13.0) | — | 112 (5.3) | — | 159 (13.9) | — | 50 (45.5) | - | 476 (23.9) | — | 2278 (71.7) | — | 10 (3.1) | — | 3229 (32.4) | — | 3229 (32.4) | — |
| Missing | — | — | — | — | — | — | - | - | — | — | — | — | — | — | 0 (0) | — | | |

* Information obtained from registers

** Response option not offered to participants

***Data only collected in long follow-ups, hence missing data independent of hospitalisation. Participants who got diagnosed at the other eight follow-ups have not answered this question

MAP-19

*1This number include higher education in Norway, encompassing both Bachelor, Master and PhD.

*2 We ask about a current psychological disorder

*3 do you want the number of COVID-19 infected at that timepoint or whether they filled out the questionnaires? Currently it is those who filled out the questionnaires at the different timepoints

Table S3: **Overview of total number of COVID-19 cases per country and proportion of population diagnosed with COVID-19**[1]

| Cohort | Location | End of study | Total cases at end of study[a] | Population of country | Proportion of population with confirmed COVID-19 diagnosis |
|---|---|---|---|---|---|
| DBDS | Denmark | August 2021 | 347 028 | 5 813 302 | 6·0% |
| EstBB-C19 | Estonia | August 2021 | 141 956 | 1 325 188 | 10·7% |
| C19-Resilience | Iceland | August 2021 | 10 789 | 343 360 | 3·1% |
| MAP-19 | Norway | August 2021 | 160 174 | 5 465 629 | 2·9% |
| MoBa | Norway | May 2021 | 125 116 | 5 465 629 | 2·3% |
| Omtanke2020 | Sweden | June 2021 | 1 089 990 | 10 160 159 | 10·7% |
| CovidLife | United Kingdom | February 2021 | 4 188 827 | 68 207 114 | 6·1% |

[a]Number of cases on the last day of the month

Table S4: **Prevalence and prevalence ratios of mental health indicators among individuals with and without a diagnosis of COVID-19, overall and in subgroup analyses by illness severity and time from diagnosis for each cohort. Combined prevalence for each mental health indicator included as well.**

| | **DBDS (DK)** N=30 917 | | | | | | | | | | | | | | | |
|---|---|---|---|---|---|---|---|---|---|---|---|---|---|---|---|---|
| | | **Overall** | | | | **Diagnosed 0-2 months ago** | | | | **Diagnosed 2-6 months ago** | | | | **Diagnosed 6-16 months ago** | | |
| | **n** | **$n_{threshold}$(%)** | **PR (age+gender adj)** | **PR (multivar adj)** [a] | **n** | **$n_{threshold}$(%)** | **PR (age+gender adj)** | **PR (multivar adj)** [a] | **n** | **$n_{threshold}$(%)** | **PR (age+gender adj)** | **PR (multivar adj)** [a] | **n** | **$n_{threshold}$(%)** | **PR (age+gender adj)** | **PR (multivar adj)** [a] |
| **Depression (PHQ-9 / EST-Q2)** | | | | | | | | | | | | | | | | |
| No confirmed diagnosis | 29 802 | 2232 (7.5) | Ref. | Ref. | - | - | - | - | - | - | - | - | - | - | - | - |
| Ever diagnosed with COVID-19 | 1111 | 136 (12.2) | 1.33 (1.13-1.56) | 1.45 (1.23-1.71) | 594 | 80 (13.5) | 1.49 (1.22-1.83) | 1.64 (1.33-2.03) | 373 | 41 (11.0) | 1.20 (0.90-1.59) | 1.28 (0.95-1.73) | 144 | 15 (10.4) | 1.06 (0.66-1.70) | 1.05 (0.65-1.70) |
| Never bedridden | 560 | 42 (7.5) | 0.81 (0.61-1.09) | 0.90 (0.67-1.22) | 311 | 27 (8.7) | 0.97 (0.68-1.38) | 1.09 (0.76-1.56) | 189 | 10 (5.3) | 0.57 (0.31-1.04) | 0.63 (0.34-1.17) | 60 | 5 (8.3) | 0.79 (0.34-1.83) | 0.82 (0.34-1.96) |
| Bedridden 1-6 days | 551 | 94 (17.1) | 1.85 (1.55-2.24) | 1.97 (1.63-2.40) | 283 | 53 (18.7) | 2.05 (1.61-2.62) | 2.22 (1.72-2.87) | 184 | 31 (16.8) | 1.85 (1.35-2.52) | 1.93 (1.38-2.67) | 84 | 10 (11.9) | 1.26 (0.71-2.24) | 1.22 (0.69-2.16) |
| Bedridden 7 days or more | - | - | - | - | - | - | - | - | - | - | - | - | - | - | - | - |
| Non-Hospitalized | - | - | - | - | - | - | - | - | - | - | - | - | - | - | - | - |
| Hospitalized | - | - | - | - | - | - | - | - | - | - | - | - | - | - | - | - |
| **Anxiety symptoms (GAD-7 / ASS / EST-Q2)** | | | | | | | | | | | | | | | | |
| No confirmed diagnosis | 29 806 | 909 (3.0) | Ref. | Ref. | - | - | - | - | - | - | - | - | - | - | - | - |
| Ever diagnosed with COVID-19 | 1111 | 39 (3.5) | 0.95 (0.96-1.30) | 0.91 (0.66-1.26) | 594 | 23 (3.9) | 1.04 (0.70-1.57) | 1.02 (0.68-1.54) | 373 | 12 (3.2) | 0.89 (0.51-1.55) | 0.82 (0.46-1.47) | 144 | <5 | - | - |
| Never bedridden | 560 | 12 (2.1) | 0.57 (0.32-1.00) | 0.55 (0.31-0.98) | 311 | 8 (2.6) | 0.69 (0.34-1.36) | 0.68 (0.34-1.36) | 189 | <5 | — | — | 60 | <5 | - | - |
| Bedridden 1-6 days | 551 | 27 (4.9) | 1.35 (0.93-1.96) | 1.29 (0.88-1.88) | 283 | 15 (5.3) | 1.45 (0.88-2.38) | 1.40 (0.85-2.30) | 184 | 9 (4.9) | 1.40 (0.73-2.65) | 1.27 (0.66-2.46) | 84 | <5 | - | - |

| | | | | | | | | | | | | | | | | |
|---|---|---|---|---|---|---|---|---|---|---|---|---|---|---|---|---|
| Bedridden 7 days or more | - | - | - | - | - | - | - | - | - | - | - | - | - | - | - | - |
| Non-Hospitalized | - | - | - | - | - | - | - | - | - | - | - | - | - | - | - | - |
| Hospitalized | - | - | - | - | - | - | - | - | - | - | - | - | - | - | - | - |
| **COVID-19 related distress symptoms (PC-PTSD-5 / PCL-5)** | | | | | | | | | | | | | | | | |
| No confirmed diagnosis | 29 806 | 8397 (28.2) | Ref. | Ref. | - | - | - | - | - | - | - | - | - | - | - | - |
| Ever diagnosed with COVID-19 | 1111 | 328 (29.5) | 1.12 (1.02-1.23) | 1.15 (1.05-1.26) | 594 | 172 (29.0) | 1.11 (0.98-1.25) | 1.14 (1.00-1.29) | 373 | 117 (31.4) | 1.17 (1.01-1.36) | 1.19 (1.03-1.38) | 144 | 39 (27.1) | 1.05 (0.81-1.36) | 1.06 (0.81-1.37) |
| Never bedridden | 560 | 151 (27.0) | 1.04 (0.91-1.19) | 1.07 (0.93-1.23) | 311 | 79 (25.4) | 0.99 (0.82-1.20) | 1.02 (0.84-1.23) | 189 | 57 (30.2) | 1.15 (0.93-1.42) | 1.18 (0.95-1.46) | 60 | 15 (25.0) | 0.96 (0.62-1.47) | 0.97 (0.63-1.50) |
| Bedridden 1-6 days | 551 | 177 (32.1) | 1.20 (1.07-1.35) | 1.22 (1.09-1.38) | 283 | 93 (32.9) | 1.23 (1.04-1.45) | 1.26 (1.07-1.48) | 184 | 60 (32.6) | 1.19 (0.97-1.46) | 1.20 (0.98-1.47) | 84 | 24 (28.6) | 1.12 (0.81-1.55) | 1.11 (0.81-1.54) |
| Bedridden 7 days or more | - | - | - | - | - | - | - | - | - | - | - | - | - | - | - | - |
| Non-Hospitalized | - | - | - | - | - | - | - | - | - | - | - | - | - | - | - | - |
| Hospitalized | - | - | - | - | - | - | - | - | - | - | - | - | - | - | - | - |
| **Sleep quality (PSQI, EST-Q2, Single-item measures)** | | | | | | | | | | | | | | | | |
| No confirmed diagnosis | - | - | - | - | - | - | - | - | - | - | - | - | - | - | - | - |
| Ever diagnosed with COVID-19 | - | - | - | - | - | - | - | - | - | - | - | - | - | - | - | - |
| Never bedridden | - | - | - | - | - | - | - | - | - | - | - | - | - | - | - | - |
| Bedridden 1-6 days | - | - | - | - | - | - | - | - | - | - | - | - | - | - | - | - |
| Bedridden 7 days or more | - | - | - | - | - | - | - | - | - | - | - | - | - | - | - | - |
| Non-Hospitalized | - | - | - | - | - | - | - | - | - | - | - | - | - | - | - | - |
| Hospitalized | - | - | - | - | - | - | - | - | - | - | - | - | - | - | - | - |

| | EstBB C-19 (EE) N=9989 | | | | | | | | | | | | | | | |
|---|---|---|---|---|---|---|---|---|---|---|---|---|---|---|---|---|
| | | Overall | | | | Diagnosed 0-2 months ago | | | | Diagnosed 2-6 months ago | | | | Diagnosed 6-16 months ago | | |
| | n | $n_{threshold}$(%) | PR (age+gender adj) | PR (multivar adj) [a] | n | $n_{threshold}$(%) | PR (age+gender adj) | PR (multivar adj) [a] | n | $n_{threshold}$(%) | PR (age+gender adj) | PR (multivar adj) [a] | n | $n_{threshold}$(%) | PR (age+gender adj) | PR (multivar adj) [a] |
| **Depression (PHQ-9 / EST-Q2)** | | | | | | | | | | | | | | | | |
| No confirmed diagnosis | 7868 | 1640 (20.8) | Ref. | Ref. | - | - | - | - | - | - | - | - | - | - | - | - |
| Ever diagnosed with COVID-19 | 2121 | 402 (19.0) | 0.92 (0.84-1.02) | 0.91 (0.83-1.00) | 1055 | 211 (20.0) | 0.96 (0.85-1.09) | 0.92 (0.82-1.04) | 954 | 171 (17.9) | 0.87 (0.76-1.01) | 0.89 (0.77-1.03) | 112 | 20 (17.9) | 0.92 (0.63-1.35) | 0.90 (0.61-1.33) |
| Never bedridden | 1061 | 174 (16.4) | 0.80 (0.70-0.92) | 0.80 (0.70-0.92) | 513 | 88 (17.2) | 0.83 (0.68-1.00) | 0.82 (0.69-0.99) | 484 | 75 (15.5) | 0.76 (0.62-0.94) | 0.76 (0.62-0.94) | 64 | 11 (17.2) | 0.94 (0.57-1.55) | 0.88 (0.53-1.46) |
| Bedridden 1-6 days | 773 | 163 (21.1) | 0.99 (0.86-1.14) | 0.96 (0.84-1.11) | 403 | 91 (22.6) | 1.05 (0.88-1.25) | 0.96 (0.81-1.15) | 339 | 66 (19.5) | 0.93 (0.74-1.15) | 0.96 (0.77-1.20) | 31 | 6 (22.9) | 0.92 (0.45-1.88) | 0.90 (0.44-1.85) |
| Bedridden 7 days or more | 287 | 65 (22.6) | 1.20 (0.97-1.49) | 1.19 (0.97-1.48) | 139 | 32 (23.0) | 1.27 (0.95-1.70) | 1.19 (0.89-1.60) | 131 | 30 (22.9) | 1.17 (0.85-1.61) | 1.22 (0.89-1.66) | 17 | 3 (17.6) | 0.86 (0.33-2.22) | 0.98 (0.34-2.80) |
| Non-Hospitalized | 2073 | 396 (19.1) | 0.93 (0.84-1.02) | 0.92 (0.83-1.01) | 1040 | 209 (20.1) | 0.97 (0.85-1.09) | 0.92 (0.82-1.04) | 931 | 168 (18.0) | 0.88 (0.76-1.01) | 0.90 (0.78-1.04) | 102 | 19 (18.6) | 0.95 (0.64-1.40) | 0.92 (0.61-1.36) |
| Hospitalized | 35 | 5 (14.3) | 0.82 (0.38-1.78) | 0.80 (0.36-1.79) | 12 | 2 (16.7) | 0.99 (0.28-3.46) | 1.08 (0.28-4.08) | 16 | 2 (12.5) | 0.73 (0.21-2.56) | 0.62 (0.18-2.07) | 7 | 1 (16.7) | 0.72 (0.14-3.65) | 0.87 (0.14-5.32) |
| **Anxiety symptoms (GAD-7 / ASS / EST-Q2)** | | | | | | | | | | | | | | | | |
| No confirmed diagnosis | 7868 | 1540 (19.6) | Ref. | Ref. | - | - | - | - | - | - | - | - | - | - | - | - |
| Ever diagnosed with COVID-19 | 2121 | 379 (17.9) | 0.93 (0.85-1.03) | 0.97 (0.87-1.07) | 1055 | 193 (18.3) | 0.95 (0.84-1.09) | 0.96 (0.84-1.09) | 954 | 164 (17.2) | 0.90 (0.78-1.03) | 0.96 (0.83-1.11) | 112 | 22 (19.6) | 1.08 (0.75-1.57) | 1.05 (0.72-1.53) |
| Never bedridden | 1061 | 174 (16.4) | 0.86 (0.75-0.99) | 0.89 (0.78-1.02) | 513 | 89 (17.3) | 0.90 (0.75-1.09) | 0.94 (0.78-1.12) | 484 | 72 (14.9) | 0.78 (0.63-0.97) | 0.81 (0.66-1.01) | 64 | 13 (20.3) | 1.18 (0.74-1.90) | 1.10 (0.67-1.80) |
| Bedridden 1-6 days | 773 | 144 (18.6) | 0.94 (0.81-1.09) | 0.97 (0.83-1.12) | 403 | 83 (20.6) | 1.03 (0.85-1.25) | 1.00 (0.83-1.21) | 339 | 57 (16.8) | 0.86 (0.68-1.08) | 0.95 (0.75-1.20) | 31 | 4 (12.9) | 0.66 (0.27-1.63) | 0.63 (0.26-1.56) |

| | | | | | | | | | | | | | | | | |
|---|---|---|---|---|---|---|---|---|---|---|---|---|---|---|---|---|
| Bedridden 7 days or more | 287 | 61 (21.3) | 1.20 (0.96-1.50) | 1.27 (1.01-1.59) | 139 | 21 (15.1) | 0.89 (0.61-1.31) | 0.92 (0.62-1.37) | 131 | 35 (26.7) | 1.44 (1.08-1.92) | 1.56 (1.18-2.08) | 17 | 5 (29.4) | 1.54 (0.77-3.09) | 1.80 (0.95-3.38) |
| Non-Hospitalized | 2073 | 372 (17.9) | 0.93 (0.85-1.03) | 0.97 (0.87-1.07) | 1040 | 192 (18.5) | 0.96 (0.84-1.09) | 0.97 (0.85-1.10) | 931 | 160 (17.2) | 0.89 (0.77-1.03) | 0.96 (0.83-1.11) | 102 | 20 (19.6) | 1.07 (0.73-1.58) | 1.03 (0.69-1.53) |
| Hospitalized | 35 | 3 (8.6) | 0.52 (0.18-1.49) | 0.53 (0.18-1.54) | 12 | 1 (8.3) | 0.51 (0.08-3.36) | 0.58 (0.08-3.99) | 16 | 1 (6.2) | 0.39 (0.06-2.59) | 0.35 (0.06-2.20) | 7 | 1 (14.3) | 0.77 (0.15-3.94) | 0.86 (0.15-5.07) |
| **COVID-19 related distress symptoms (PC-PTSD-5 / PCL-5)** | | | | | | | | | | | | | | | | |
| No confirmed diagnosis | - | - | - | - | - | - | - | - | - | - | - | - | - | - | - | - |
| Ever diagnosed with COVID-19 | - | - | - | - | - | - | - | - | - | - | - | - | - | - | - | - |
| Never bedridden | - | - | - | - | - | - | - | - | - | - | - | - | - | - | - | - |
| Bedridden 1-6 days | - | - | - | - | - | - | - | - | - | - | - | - | - | - | - | - |
| Bedridden 7 days or more | - | - | - | - | - | - | - | - | - | - | - | - | - | - | - | - |
| Non-Hospitalized | - | - | - | - | - | - | - | - | - | - | - | - | - | - | - | - |
| Hospitalized | - | - | - | - | - | - | - | - | - | - | - | - | - | - | - | - |
| **Sleep quality (PSQI, EST-Q2, Single-item measures)** | | | | | | | | | | | | | | | | |
| No confirmed diagnosis | 7868 | 2406 (30.6) | Ref. | Ref. | - | - | - | - | - | - | - | - | - | - | - | - |
| Ever diagnosed with COVID-19 | 2121 | 642 (30.3) | 1.01 (0.94-1.08) | 1.03 (0.96-1.11) | 1055 | 304 (28.8) | 0.97 (0.88-1.08) | 0.98 (0.89-1.09) | 954 | 295 (30.9) | 1.02 (0.92-1.12) | 1.07 (0.97-1.18) | 112 | 43 (38.4) | 1.26 (0.99-1.60) | 1.19 (0.94-1.52) |
| Never bedridden | 1061 | 319 (30.1) | 1.00 (0.91-1.10) | 1.02 (0.93-1.12) | 513 | 142 (27.7) | 0.94 (0.81-1.08) | 0.96 (0.84-1.10) | 484 | 151 (31.2) | 1.03 (0.90-1.17) | 1.05 (0.92-1.20) | 64 | 26 (40.6) | 1.30 (0.96-1.76) | 1.24 (0.91-1.70) |
| Bedridden 1-6 days | 773 | 223 (28.8) | 0.98 (0.87-1.09) | 0.99 (0.88-1.11) | 403 | 117 (29.0) | 0.99 (0.85-1.16) | 0.97 (0.83-1.14) | 339 | 94 (27.7) | 0.93 (0.78-1.10) | 0.99 (0.83-1.18) | 31 | 12 (38.7) | 1.31 (0.85-2.03) | 1.17 (0.75-1.82) |
| Bedridden 7 days or more | 287 | 100 (34.8) | 1.13 (0.96-1.32) | 1.19 (1.01-1.39) | 139 | 45 (32.4) | 1.06 (0.83-1.35) | 1.10 (0.86-1.41) | 131 | 50 (38.2) | 1.21 (0.97-1.49) | 1.30 (1.06-1.60) | 17 | 5 (29.4) | 1.01 (0.48-2.11) | 1.04 (0.51-2.10) |

| | | | | | | | | | | | | | | | | |
|---|---|---|---|---|---|---|---|---|---|---|---|---|---|---|---|---|
| Non-Hospitalized | 2073 | 628 (30.3) | 1.01 (0.94-1.09) | 1.04 (0.96-1.11) | 1040 | 299 (28.7) | 0.97 (0.88-1.08) | 0.98 (0.89-1.08) | 931 | 288 (30.9) | 1.02 (0.92-1.13) | 1.07 (0.97-1.19) | 102 | 41 (40.2) | 1.32 (1.04-1.67) | 1.24 (0.97-1.59) |
| Hospitalized | 35 | 10 (28.6) | 0.89 (0.53-1.48) | 0.91 (0.56-1.47) | 12 | 4 (33.3) | 1.02 (0.48-2.19) | 1.18 (0.55-2.51) | 16 | 4 (25.0) | 0.76 (0.33-1.74) | 0.73 (0.33-1.60) | 7 | 2 (28.6) | 0.98 (0.31-3.02) | 0.95 (0.37-2.45) |

| | Iceland N=21 615 | | | | | | | | | | | | | | | |
|---|---|---|---|---|---|---|---|---|---|---|---|---|---|---|---|---|
| | | **Overall** | | | | **Diagnosed 0-2 months ago** | | | | **Diagnosed 2-6 months ago** | | | | **Diagnosed 6-16 months ago** | | |
| | **n** | **$n_{threshold}$(%)** | **PR (age+gender adj)** | **PR (multivar adj) [a]** | **n** | **$n_{threshold}$(%)** | **PR (age+gender adj)** | **PR (multivar adj) [a]** | **n** | **$n_{threshold}$(%)** | **PR (age+gender adj)** | **PR (multivar adj) [a]** | **n** | **$n_{threshold}$ (%)** | **PR (age+gender adj)** | **PR (multivar adj) [a]** |
| **Depression (PHQ-9 / EST-Q2)** | | | | | | | | | | | | | | | | |
| No confirmed diagnosis | 20 471 | 3548 (17.3) | Ref. | Ref. | - | - | - | - | - | - | - | - | - | - | - | - |
| Ever diagnosed with COVID-19 | 1144 | 242 (21.2) | 1.02 (0.91-1.14) | 1.19 (1.01-1.39) | 444 | 103 (23.2) | 1.13 (0.95-1.34) | 1.21 (1.02-1.45) | 541 | 122 (22.6) | 1.03 (0.89-1.21) | 1.08 (0.81-1.45) | 159 | 17 (10.7) | 0.59 (0.38-0.92) | 0.84 (0.43-1.65) |
| Never bedridden | 428 | 57 (13.3) | 0.60 (0.47-0.76) | 0.75 (0.58-0.97) | 159 | 19 (11.9 | 0.54 (0.35-0.82) | 0.66 (0.43-0.99) | 206 | 33 (16.0) | 0.70 (0.51-0.95) | 0.78 (0.52-1.15) | 63 | 5 (7.9) | 0.40 (0.18-0.91) | 0.58 (0.22-1.52) |
| Bedridden 1-6 days | 455 | 102 (22.4) | 1.05 (0.89-1.24) | 1.12 (0.91-1.37) | 165 | 43 (26.1) | 1.27 (0.99-1.62) | 1.18 (0.91-1.53) | 236 | 53 (22.5) | 1.02 (0.81-1.27) | 0.98 (0.70-1.37) | 54 | 6 (11.1) | 0.56 (0.26-1.21) | 0.94 (0.42-2.14) |
| Bedridden 7 days or more | 261 | 83 (31.8) | 1.79 (1.51-2.12) | 1.95 (1.60-2.38) | 120 | 41 (34.2) | 1.87 (1.48-2.38) | 2.06 (1.64-2.60) | 99 | 36 (36.4) | 1.93 (1.51-2.47) | 1.82 (1.26-2.63) | 42 | 6 (14.3) | 1.03 (0.51-2.09) | 1.17 (0.47-2.88) |
| Non-Hospitalized | 1056 | 214 (20.3) | 0.95 (0.84-1.07) | 1.10 (0.93-1.29) | 407 | 89 (21.9) | 1.04 (0.86-1.24) | 1.13 (0.94-1.36) | 505 | 112 (22.2) | 0.99 (0.84-1.16) | 1.03 (0.77-1.38) | 144 | 13 (9.0) | 0.47 (0.28-0.78) | 0.68 (0.35-1.35) |
| Hospitalized | 61 | 19 (31.1) | 2.20 (1.54-3.15) | 2.17 (1.44-3.26) | 22 | 10 (45.5) | 2.75 (1.90-3.99) | 2.27 (1.36-3.81) | 30 | 6 (20.0) | 1.44 (0.68-3.05) | 1.56 (0.69-3.53) | 9 | 3 (33.3) | 3.51 (1.34-9.16) | 3.64 (1.64-8.09) |
| **Anxiety symptoms (GAD-7 / ASS / EST-Q2)** | | | | | | | | | | | | | | | | |
| No confirmed diagnosis | 20 471 | 2365 (11.6) | Ref. | Ref. | - | - | - | - | - | - | - | - | - | - | - | - |

| | | | | | | | | | | | | | | | | |
|---|---|---|---|---|---|---|---|---|---|---|---|---|---|---|---|---|
| Ever diagnosed with COVID-19 | 1144 | 132 (11.5) | 0.80 (0.68-0.94) | 1.01 (0.81-1.26) | 444 | 66 (14.9) | 1.05 (0.84-1.31) | 1.13 (0.90-1.42) | 541 | 62 (11.5) | 0.75 (0.59-0.94) | 0.77 (0.51-1.15) | 159 | 4 (2.5) | 0.20 (0.08-0.53) | 0.21 (0.07-0.65) |
| Never bedridden | 428 | 31 (7.2) | 0.46 (0.33-0.65) | 0.64 (0.43-0.94) | 159 | 18 (11.3) | 0.72 (0.47-1.11) | 0.89 (0.58-1.36) | 206 | 13 (6.3) | 0.39 (0.23-0.65) | 0.44 (0.23-0.86) | 63 | 0 (0) | - | - |
| Bedridden 1-6 days | 455 | 60 (13.2) | 0.89 (0.71-1.12) | 1.02 (0.78-1.34) | 165 | 22 (13.3) | 0.94 (0.65-1.38) | 0.86 (0.59-1.25) | 236 | 36 (15.3) | 0.99 (0.74-1.32) | 0.95 (0.60-1.50) | 54 | 2 (3.7) | 0.27 (0.07-1.04) | 0.38 (0.09-1.52) |
| Bedridden 7 days or more | 261 | 41 (15.7) | 1.31 (1.00-1.72) | 1.57 (1.17-2.12) | 120 | 26 (21.7) | 1.76 (1.26-2.46) | 1.99 (1.43-2.76) | 99 | 13 (13.1) | 1.02 (0.63-1.64) | 0.99 (0.57-1.73) | 42 | 2 (4.8) | 0.54 (0.15-1.92) | 0.40 (0.11-1.55) |
| Non-Hospitalized | 1056 | 121 (11.5) | 0.77 (0.65-0.91) | 0.97 (0.77-1.22) | 407 | 59 (14.5) | 0.99 (0.78-1.26) | 1.08 (0.85-1.38) | 505 | 58 (11.5) | 0.73 (0.57-0.92) | 0.73 (0.48-1.12) | 144 | 4 (2.8) | 0.21 (0.08-0.55) | 0.23 (0.07-0.69) |
| Hospitalized | 61 | 10 (16.4) | 1.81 (1.10-2.96) | 1.82 (1.07-3.10) | 22 | 6 (27.3) | 2.45 (1.50-4.02) | 1.93 (1.08-3.45) | 30 | 4 (13.3) | 1.53 (0.62-3.80) | 1.57 (0.58-4.28) | 9 | 0 (0) | - | - |
| **COVID-19 related distress symptoms (PC-PTSD-5 / PCL-5)** | | | | | | | | | | | | | | | | |
| No confirmed diagnosis | 20 471 | 3902 (19.1) | Ref. | Ref. | - | - | - | - | - | - | - | - | - | - | - | - |
| Ever diagnosed with COVID-19 | 1144 | 223 (19.5) | 0.93 (0.83-1.05) | 1.14 (0.97-1.35) | 444 | 108 (24.3) | 1.16 (0.98-1.36) | 1.23 (1.04-1.46) | 541 | 105 (19.4) | 0.9 (0.76-1.07) | 0.86 (0.62-1.20) | 159 | 10 (6.3) | 0.34 (0.19-0.62) | 0.36 (0.16-0.82) |
| Never bedridden | 428 | 49 (11.4) | 0.53 (0.41-0.69) | 0.67 (0.50-0.90) | 159 | 27 (17.0) | 0.79 (0.56-1.10) | 0.89 (0.63-1.25) | 206 | 20 (9.7) | 0.44 (0.29-0.67) | 0.44 (0.26-0.72) | 63 | 2 (3.2) | 0.16 (0.04-0.64) | 0.17 (0.04-0.73) |
| Bedridden 1-6 days | 455 | 99 (21.8) | 1.02 (0.85-1.21) | 1.20 (0.98-1.48) | 165 | 39 (23.6) | 1.10 (0.84-1.45) | 1.11 (0.84-1.47) | 236 | 56 (23.7) | 1.09 (0.87-1.36) | 1.01 (0.70-1.47) | 54 | 4 (7.4) | 0.38 (0.15-0.97) | 0.47 (0.16-1.33) |
| Bedridden 7 days or more | 261 | 75 (28.7) | 1.51 (1.25-1.82) | 1.73 (1.40-2.14) | 120 | 42 (35.0) | 1.77 (1.40-2.25) | 1.89 (1.48-2.41) | 99 | 29 (29.3) | 1.49 (1.09-2.04) | 1.35 (0.90-2.04) | 42 | 4 (9.5) | 0.61 (0.25-1.49) | 0.54 (0.17-1.73) |
| Non-Hospitalized | 1056 | 199 (18.8) | 0.88 (0.78-1.00) | 1.08 (0.91-1.28) | 407 | 96 (23.6) | 1.10 (0.92-1.31) | 1.18 (0.99-1.42) | 505 | 94 (18.6) | 0.85 (0.71-1.02) | 0.81 (0.58-1.13) | 144 | 9 (6.2) | 0.32 (0.17-0.61) | 0.35 (0.16-0.78) |
| Hospitalized | 61 | 17 (27.9) | 1.75 (1.17-2.60) | 1.92 (1.28-2.88) | 22 | 9 (40.9) | 2.23 (1.40-3.57) | 2.09 (1.25-3.49) | 30 | 7 (23.3) | 1.51 (0.77-2.96) | 1.43 (0.73-2.82) | 9 | 1 (11.1) | 0.94 (0.14-6.17) | 0.81 (0.10-6.72) |
| **Sleep quality (PSQI, EST-Q2, Single-item measures)** | | | | | | | | | | | | | | | | |

| | | | | | | | | | | | | | | | | |
|---|---|---|---|---|---|---|---|---|---|---|---|---|---|---|---|---|
| No confirmed diagnosis | 20 471 | 5578 (27.2) | Ref. | Ref. | - | - | - | - | - | - | - | - | - | - | - | - |
| Ever diagnosed with COVID-19 | 1144 | 393 (34.4) | 1.15 (1.05-1.25) | 1.09 (0.97-1.24) | 444 | 144 (32.4) | 1.08 (0.95-1.24) | 1.08 (0.94-1.24) | 541 | 206 (38.1) | 1.24 (1.11-1.38) | 1.04 (0.83-1.30) | 159 | 43 (27.0) | 0.97 (0.75-1.25) | 0.74 (0.51-1.06) |
| Never bedridden | 428 | 127 (29.7) | 0.95 (0.82-1.11) | 0.94 (0.79-1.11) | 159 | 43 (27.0) | 0.87 (0.67-1.12) | 0.92 (0.71-1.18) | 206 | 63 (30.6) | 0.96 (0.78-1.19) | 0.83 (0.62-1.11) | 63 | 21 (33.3) | 1.14 (0.81-1.61) | 0.90 (0.58-1.38) |
| Bedridden 1-6 days | 455 | 154 (33.8) | 1.11 (0.98-1.27) | 1.02 (0.87-1.19) | 165 | 51 (30.9) | 1.03 (0.82-1.29) | 0.96 (0.76-1.20) | 236 | 92 (39.0) | 1.26 (1.07-1.47) | 1.03 (0.79-1.33) | 54 | 11 (20.4) | 0.70 (0.42-1.17) | 0.56 (0.32-0.99) |
| Bedridden 7 days or more | 261 | 112 (42.9) | 1.55 (1.35-1.79) | 1.44 (1.22-1.69) | 120 | 50 (41.7) | 1.49 (1.21-1.83) | 1.49 (1.22-1.83) | 99 | 51 (51.5) | 1.81 (1.49-2.21) | 1.45 (1.10-1.91) | 42 | 11 (26.2) | 1.07 (0.64-1.77) | 0.72 (0.40-1.28) |
| Non-Hospitalized | 1056 | 356 (33.7) | 1.10 (1.01-1.21) | 1.06 (0.94-1.20) | 407 | 130 (31.9) | 1.05 (0.91-1.21) | 1.06 (0.91-1.23) | 505 | 187 (37.0) | 1.19 (1.06-1.33) | 1.00 (0.79-1.25) | 144 | 39 (27.1) | 0.94 (0.72-1.23) | 0.73 (0.51-1.05) |
| Hospitalized | 61 | 29 (47.5) | 1.93 (1.51-2.48) | 1.66 (1.27-2.17) | 22 | 10 (45.5) | 1.74 (1.18-2.55) | 1.47 (1.00-2.17) | 30 | 16 (53.3) | 2.16 (1.55-3.00) | 1.74 (1.17-2.60) | 9 | 3 (33.3) | 1.63 (0.63-4.18) | 1.10 (0.40-2.98) |

| | MAP-19 (NO) N=10 061 | | | | | | | | | | | | | | | |
|---|---|---|---|---|---|---|---|---|---|---|---|---|---|---|---|---|
| | | **Overall** | | | | **Diagnosed 0-2 months ago** | | | | **Diagnosed 2-6 months ago** | | | | **Diagnosed 6-16 months ago** | | |
| | **n** | **$n_{threshold}$(%)** | **PR (age+gender adj)** | **PR (multivar adj)** [a] | **n** | **$n_{threshold}$(%)** | **PR (age+gender adj)** | **PR (multivar adj)** [a] | **n** | **$n_{threshold}$(%)** | **PR (age+gender adj)** | **PR (multivar adj)** [a] | **n** | **$n_{threshold}$(%)** | **PR (age+gender adj)** | **PR (multivar adj)** [a] |
| **Depression (PHQ-9 / EST-Q2)** | | | | | | | | | | | | | | | | |
| No confirmed diagnosis | 9951 | 3064 (30.8) | Ref. | Ref. | | | | | | | | | | | | |
| Ever diagnosed with COVID-19 | 110 | 38 (34.5) | 1.06 (0.81 - 1.37) | 1.09 (0.83-1.45) | 17 | 7 (41.2) | 1.29 (0.70-2.38) | 1.56 (0.82-2.98) | 43 | 18 (41.9) | 1.24 (0.85-1.80) | 1.20 (0.80-1.80) | 50 | 13 (26) | 0.81 (0.51-1.28) | 0.86 (0.54-1.38) |
| Never bedridden | - | - | - | - | - | - | - | - | - | - | - | - | - | - | - | - |
| Bedridden 1-6 days | - | - | - | - | - | - | - | - | - | - | - | - | - | - | - | - |
| Bedridden 7 days or more | - | - | - | - | - | - | - | - | - | - | - | - | - | - | - | - |
| Non-Hospitalized | - | - | - | - | - | - | - | - | - | - | - | - | - | - | - | - |

| | | | | | | | | | | | | | | | | |
|---|---|---|---|---|---|---|---|---|---|---|---|---|---|---|---|---|
| Hospitalized | - | - | - | - | - | - | - | - | - | - | - | - | - | - | - | - |
| **Anxiety symptoms (GAD-7 / ASS / EST-Q2)** | | | | | | | | | | | | | | | | |
| No confirmed diagnosis | 9951 | 1850 (18.6) | Ref. | Ref | | | | | | | | | | | | |
| Ever diagnosed with COVID-19 | 110 | 17 (15.5) | 0.78 (0.51-1.20) | 0.81 (0.53-1.23) | 17 | 3 (17.6) | 0.91 (0.32-2.49) | 1.16 (0.42-3.21) | 43 | 6 (14.0) | 0.69 (0.89-1.42) | 0.65 (0.32-1.32) | 50 | 8 (16.0) | 0.82 (0.43-1.53) | 0.87 (0.48-1.58) |
| Never bedridden | - | - | - | - | - | - | - | - | - | - | - | - | - | - | - | - |
| Bedridden 1-6 days | - | - | - | - | - | - | - | - | - | - | - | - | - | - | - | - |
| Bedridden 7 days or more | - | - | - | - | - | - | - | - | - | - | - | - | - | - | - | - |
| Non-Hospitalized | - | - | - | - | - | - | - | - | - | - | - | - | - | - | - | - |
| Hospitalized | - | - | - | - | - | - | - | - | - | - | - | - | - | - | - | - |
| **COVID-19 related distress symptoms (PC-PTSD-5 / PCL-5)** | | | | | | | | | | | | | | | | |
| No confirmed diagnosis | 4491 | 858 (8.6) | Ref | Ref | | | | | | | | | | | | |
| Ever diagnosed with COVID-19 | 110 | 19 (17.3) | 0.89 (0.59-1.35) | 0.89 (0.59-1.34) | 17 | 3 (17.6) | 0.92 (0.33-2.57) | 0.95 (0.35-2.63) | 43 | 8 (18.6) | 0.96 (0.51-1.79) | 0.94 (0.50-1.77) | 50 | 8 (16.0) | 0.83 (0.44-1.58) | 0.83 (0.45-1.55) |
| Never bedridden | - | - | - | - | - | - | - | - | - | - | - | - | - | - | - | - |
| Bedridden 1-6 days | - | - | - | - | - | - | - | - | - | - | - | - | - | - | - | - |
| Bedridden 7 days or more | - | - | - | - | - | - | - | - | - | - | - | - | - | - | - | - |
| Non-Hospitalized | - | - | - | - | - | - | - | - | - | - | - | - | - | - | - | - |
| Hospitalized | - | - | - | - | - | - | - | - | - | - | - | - | - | - | - | - |
| **Sleep quality (PSQI, EST-Q2, Single-item measures)** | | | | | | | | | | | | | | | | |
| No confirmed diagnosis | - | - | - | - | - | - | - | - | - | - | - | - | - | - | - | - |

| | | | | | | | | | | | | | | | | |
|---|---|---|---|---|---|---|---|---|---|---|---|---|---|---|---|---|
| Ever diagnosed with COVID-19 | - | - | - | - | - | - | - | - | - | - | - | - | - | - | - | - |
| Never bedridden | - | - | - | - | - | - | - | - | - | - | - | - | - | - | - | - |
| Bedridden 1-6 days | - | - | - | - | - | - | - | - | - | - | - | - | - | - | - | - |
| Bedridden 7 days or more | - | - | - | - | - | - | - | - | - | - | - | - | - | - | - | - |
| Non-Hospitalized | - | - | - | - | - | - | - | - | - | - | - | - | - | - | - | - |
| Hospitalized | - | - | - | - | - | - | - | - | - | - | - | - | - | - | - | - |

| | MoBa (NO) N=132 451 | | | | | | | | | | | | | | | |
|---|---|---|---|---|---|---|---|---|---|---|---|---|---|---|---|---|
| | | Overall | | | | Diagnosed 0-2 months ago | | | | Diagnosed 2-6 months ago | | | | Diagnosed 6-16 months ago | | |
| | **n** | **$n_{threshold}$(%)** | **PR (age+gender adj)** | **PR (multivar adj)** [a] | **n** | **$n_{threshold}$(%)** | **PR (age+gender adj)** | **PR (multivar adj)** [a] | **n** | **$n_{threshold}$(%)** | **PR (age+gender adj)** | **PR (multivar adj)** [a] | **n** | **$n_{threshold}$ (%)** | **PR (age+gender adj)** | **PR (multivar adj)** [a] |
| **Depression (PHQ-9 / EST-Q2)** | | | | | | | | | | | | | | | | |
| No confirmed diagnosis | 91 649 | 4463 (4.9) | Ref | Ref | - | - | - | - | - | - | - | - | - | - | - | - |
| Ever diagnosed with COVID-19 | 292 | 19 (6.5) | 1.40 (0.91-2.17) | 1.46 (0.94-2.26) | 23 | 3 (13.0) | 2.61 (0.92-7.38) | 1.99 (0.72-5.45) | 251 | 14 (5.6) | 1.21 (0.73-2.02) | 1.29 (0.77-2.17) | - | - | - | - |
| Never bedridden | 85 | 3 (3.5) | 0.77 (0.25-2.33) | 0.84 (0.27-2.63) | 8 | 0 (0) | - | - | 77 | 3 (3.9) | 0.85 (0.28-2.57) | 0,96 (0.31-2.99) | - | - | - | - |
| Bedridden 1-6 days | 18 | 0 (0) | - | - | 0 | 0 (0) | - | - | 18 | 0 (0) | - | - | - | - | - | - |
| Bedridden 7 days or more | 115 | 9 (7.8) | 1.64 (0.88-3.06) | 1.87 (0.99-3.51) | 4 | 1 (25.0) | 4.30 (0.78-23.59) | 4.46 (0.77-25.73) | 111 | 8 (7.2) | 1.52 (0.78-2.96) | 1.74 (0.89-3.41) | - | - | - | - |
| Non-Hospitalized | 211 | 12 (5.7) | 1.21 (0.70-2.10) | 1.35 (0.77-2.36) | 12 | 1 (8.3) | 1.65 (0.26-10.58) | 1.51 (0.22-10.32) | 199 | 11 (5.5) | 1.18 (0.67-2.10) | 1.34 (0.75-2.40) | - | - | - | - |
| Hospitalized | 7 | 0 (0) | - | - | 0 | 0 (0) | - | - | 7 | 0 (0) | - | - | - | - | - | - |
| **Anxiety symptoms** | | | | | | | | | | | | | | | | |

| | | | | | | | | | | | | | | | | |
|---|---|---|---|---|---|---|---|---|---|---|---|---|---|---|---|---|
| **(GAD-7 / ASS / EST-Q2)** | | | | | | | | | | | | | | | | |
| No confirmed diagnosis | 98 195 | 2724 (2.8) | Ref. | Ref. | - | - | - | - | - | - | - | - | - | - | - | - |
| Ever diagnosed with COVID-19 | 295 | 10 (3.4) | 1.26 (0.68-2.31) | 1.26 (0.69-2.32) | 102 | 2 (2.0) | 0.72 (0.18-2.87) | 0.59 (0.15-2.28) | 193 | 8 (4.1) | 1.54 (0.78-3.03) | 1.77 (0.89-3.50) | - | - | - | - |
| Never bedridden | 79 | 3 (3.8) | 1.43 (0.47-4.34) | 1.64 (0.58-4.63) | 10 | 1 (10.0) | 4.22 (0.66-26.99) | 4.02 (1.02-15.78) | 69 | 2 (2.9) | 1.07 (0.27-4.21) | 1.27 (0.34-4.72) | - | - | - | - |
| Bedridden 1-6 days | 19 | 0 (0) | - | - | 2 | 0 (0) | - | - | 17 | 0 (0) | - | - | - | - | - | - |
| Bedridden 7 days or more | 122 | 6 (4.9) | 1.81 (0.83-3.94) | 2.07 (0.93-4.6) | 15 | 0 (0) | - | - | 107 | 6 (5.6) | 2.08 (0.96-4.52) | 2.39 (1.08-5.32) | - | - | - | - |
| Non-Hospitalized | 212 | 9 (4.2) | 1.57 (0.83-2.98) | 1.78 (0.95-3.37) | 23 | 1 (4.3) | 1.57 (0.23-10.77) | 1.67 (0.30-9.36) | 189 | 8 (4.2) | 1.57 (0.80-3.09) | 1.8 (0.91-3.57) | - | - | - | - |
| Hospitalized | 8 | 0 (0.0) | - | - | 4 | 0 (0) | - | - | 4 | 0 (0) | - | - | - | - | - | - |
| **COVID-19 related distress symptoms (PC-PTSD-5 / PCL-5)** | | | | | | | | | | | | | | | | |
| No confirmed diagnosis | - | - | - | - | - | - | - | - | - | - | - | - | - | - | - | - |
| Ever diagnosed with COVID-19 | - | - | - | - | - | - | - | - | - | - | - | - | - | - | - | - |
| Never bedridden | - | - | - | - | - | - | - | - | - | - | - | - | - | - | - | - |
| Bedridden 1-6 days | - | - | - | - | - | - | - | - | - | - | - | - | - | - | - | - |
| Bedridden 7 days or more | - | - | - | - | - | - | - | - | - | - | - | - | - | - | - | - |
| Non-Hospitalized | - | - | - | - | - | - | - | - | - | - | - | - | - | - | - | - |
| Hospitalized | - | - | - | - | - | - | - | - | - | - | - | - | - | - | - | - |
| **Sleep quality (PSQI, EST-Q2, Single-item measures)** | | | | | | | | | | | | | | | | |
| No confirmed diagnosis | 77 044 | 16 062 (20.8) | Ref. | Ref. | - | - | - | - | - | - | - | - | - | - | - | - |
| Ever diagnosed with COVID-19 | 1061 | 289 (27.2) | 1.32 (1.19-1.46) | 1.32 (1.20-1.46) | 205 | 49 (23.9) | 1.15 (0.90-1.47) | 1.15 (0.90-1.47) | 568 | 162 (28.5) | 1.38 (1.21-1.57) | 1.36 (1.20-1.55) | 288 | 78 (27.1) | 1.32 (1.09-1.60) | 1.35 (1.12-1.63) |

| | n | $n_{threshold}$ (%) | PR (age+gender adj) | PR (multivar adj) [a] | n | $n_{threshold}$ (%) | PR (age+gender adj) | PR (multivar adj) [a] | n | $n_{threshold}$ (%) | PR (age+gender adj) | PR (multivar adj) [a] | n | $n_{threshold}$ (%) | PR (age+gender adj) | PR (multivar adj) [a] |
|---|---|---|---|---|---|---|---|---|---|---|---|---|---|---|---|---|
| Never bedridden | 398 | 80 (20.1) | 0.98 (0.81-1.20) | 1.01 (0.83-1.23) | 62 | 12 (19.4) | 0.94 (0.57-1.56) | 0.94 (0.57-1.55) | 243 | 52 (21.4) | 1.04 (0.82-1.33) | 1.07 (0.84-1.36) | 93 | 16 (17.2) | 0.86 (0.55-1.35) | 0.91 (0.58-1.43) |
| Bedridden 1-6 days | 124 | 31 (25.0) | 1.24 (0.92-1.68) | 1.24 (0.92-1.67) | 46 | 10 (21.7) | 1.07 (0.62-1.85) | 1.09 (0.64-1.87) | 61 | 18 (29.5) | 1.46 (0.99-2.14) | 1.43 (0.98-2.08) | 17 | 3 (17.6) | 0.91 (0.32-2.53) | 0.95 (0.36-2.48) |
| Bedridden 7 days or more | 410 | 145 (35.4) | 1.68 (1.47-1.91) | 1.64 (1.44-1.87) | 61 | 18 (29.5) | 1.38 (0.94-2.04) | 1.35 (0.91-2.00) | 215 | 80 (37.2) | 1.75 (1.47-2.08) | 1.69 (1.42-2.01) | 134 | 47 (35.1) | 1.69 (1.34-2.12) | 1.70 (1.35-2.13) |
| Non-Hospitalized | 912 | 249 (27.3) | 1.32 (1.18-1.47) | 1.32 (1.19-1.47) | 168 | 39 (23.2) | 1.12 (0.85-1.47) | 1.11 (0.85-1.46) | 510 | 149 (29.2) | 1.40 (1.22-1.61) | 1.40 (1.22-1.60) | 234 | 61 (26.1) | 1.27 (1.03-1.58) | 1.32 (1.06-1.63) |
| Hospitalized | 20 | 7 (35.0) | 1.85 (1.03-3.30) | 1.71 (0.95-3.07) | 1 | 1 (100.0) | 5.98 (5.83-6.14) | 6.33 (6.01-6.67) | 9 | 1 (11.1) | 0.60 (0.10-3.63) | 0.58 (0.10-3.36) | 10 | 5 (50.0) | 2.54 (1.38-4.66) | 2.25 (1.17-4.3) |

| | Omtanke (SE) N=23 698 | | | | | | | | | | | | | | | |
|---|---|---|---|---|---|---|---|---|---|---|---|---|---|---|---|---|
| | | Overall | | | | Diagnosed 0-2 months ago | | | | Diagnosed 2-6 months ago | | | | Diagnosed 6-16 months ago | | |
| | n | $n_{threshold}$ (%) | PR (age+gender adj) | PR (multivar adj) [a] | n | $n_{threshold}$ (%) | PR (age+gender adj) | PR (multivar adj) [a] | n | $n_{threshold}$ (%) | PR (age+gender adj) | PR (multivar adj) [a] | n | $n_{threshold}$ (%) | PR (age+gender adj) | PR (multivar adj) [a] |
| **Depression (PHQ-9 / EST-Q2)** | | | | | | | | | | | | | | | | |
| No confirmed diagnosis | 20 010 | 3702 (18,5) | Ref. | Ref. | - | - | - | - | - | - | - | - | - | - | - | - |
| Ever diagnosed with COVID-19 | 3060 | 706 (23.1) | 1.16 (1.09 - 1.25) | 1.18 (1.10 - 1.27) | 209 | 48 (23.0) | 1.42 (1.07 - 1.89) | 1.42 (1.00 - 2.02) | 933 | 219 (23.5) | 1.10 (0.97 - 1.24) | 1.09 (0.97 - 1.23) | 1918 | 439 (22.9) | 1.17 (1.07 - 1.28) | 1.26 (1.15 - 1.38) |
| Never bedridden | 451 | 77 (17,1) | 0.80 (0.65 - 0.98) | 0.90 (0.74 - 1.09) | 10 | 2 (20.0) | 1.17 (0.32 - 4.28) | 0.87 (0.24 - 3.13) | 207 | 34 (16.4) | 0.72 (0.53 - 0.98) | 0.77 (0.56 - 1.04) | 234 | 44 (17.5) | 0.86 (0.648 - 1.13) | 1.04 (0.79 - 1.35) |
| Bedridden 1-6 days | 433 | 93 (21.5) | 1.03 (0.87 - 1.23) | 1.05 (0.88 - 1.26) | 15 | 2 (13.3) | 0.96 (0.26 - 3.31) | 0.95 (0.270 - 3.35) | 184 | 47 (25.5) | 1.17 (0.92 - 1.50) | 1.21 (0.95 - 1.53) | 234 | 41 (18.8) | 0.91 (0.70 - 1.17) | 0.94 (0.72 - 1.23) |
| Bedridden 7 days or more | 367 | 142 (38.7) | 2.12 (1.87 - 2.40) | 1.69 (1.47 - 1.93) | 9 | 3 (33.3) | 2.05 (0.83 - 5.09) | 1.51 (0.62 - 3.70) | 97 | 38 (39.2) | 2.09 (1.64 - 2.66) | 1.60 (1.22 - 2.09) | 261 | 101 (38.7) | 2.09 (1.80 - 2.43) | 1.75 (1.49 - 2.05) |
| Non-Hospitalized | 2978 | 671 (22.5) | 1.12 (1.05 - 1.21) | 1.16 (1.08 - 1.24) | 208 | 48 (23.1) | 1.43 (1.08 - 1.89) | 1.43 (1.01 - 2.02) | 918 | 214 (23.3) | 1.08 (0.95 - 1.22) | 1.09 (0.96 - 1.22) | 1852 | 409 (22.1) | 1.11 (1.01 - 1.22) | 1.23 (1.12 - 1.35) |

| | | | | | | | | | | | | | | | | |
|---|---|---|---|---|---|---|---|---|---|---|---|---|---|---|---|---|
| Hospitalized | 63 | 28 (44.4) | 2.67 (2.08 - 3.43) | 1.67 (1.24 - 2.25) | 1 | 0 (0) | - | - | 12 | 3 (25.0) | 2.75 (1.24 - 6.10) | 1.44 (0.56 - 3.71) | 50 | 25 (50.0) | 2.59 (1.99 - 3.36) | 1.76 (1.29 - 2.40) |
| **Anxiety symptoms (GAD-7 / ASS / EST-Q2)** | | | | | | | | | | | | | | | | |
| No confirmed diagnosis | 20 327 | 2755 (13.6) | Ref. | Ref. | - | - | - | - | - | - | - | - | - | - | - | - |
| Ever diagnosed with COVID-19 | 3154 | 429 (13.6) | 0.93 (0.85 - 1.02) | 0.95 (0.87 - 1.05) | 212 | 24 (11.3) | 0.91 (0.61 - 1.36) | 1.04 (0.66 - 1.65) | 958 | 125 (13.0) | 0.85 (0.72 - 1.01) | 0.85 (0.71 - 1.00) | 1984 | 280 (14.1) | 0.94 (0.84 - 1.06) | 1.03 (0.92 - 1.16) |
| Never bedridden | 474 | 47 (9.9) | 0.64 (0.49 - 0.88) | 0.71 (0.55 - 0.92) | 12 | 2 (16.7) | 1.28 (0.33 - 5.00) | 1.02 (0.31 - 3.34) | 211 | 20 (9.5) | 0.61 (0.40 - 0.92) | 0.64 (0.43 - 0.96) | 251 | 25 (10.0) | 0.63 (0.44 - 0.91) | 0.74 (0.52 - 1.06) |
| Bedridden 1-6 days | 459 | 69 (15.0) | 1.01 (0.82 - 1.25) | 1.03 (0.84 - 1.28) | 15 | 2 (13.3) | 1.53 (0.42 - 5.60) | 1.51 (0.37 - 6.14) | 193 | 33 (17.1) | 1.15 (0.85 - 1.55) | 1.16 (0.86 - 1.56) | 251 | 34 (13.5) | 0.87 (0.64 - 1.18) | 0.91 (0.67 - 1.24) |
| Bedridden 7 days or more | 377 | 86 (22.8) | 1.78 (1.48 - 2.13) | 1.46 (1.21 - 1.77) | 9 | 1 (11.1) | 1.01 (0.16 - 6.18) | 0.76 (0.17 - 3.46) | 99 | 23 (23.2) | 1.81 (1.31 - 2.49) | 1.43 (1.00 - 2.06) | 269 | 62 (23.0) | 1.72 (1.38 - 2.14) | 1.49 (1.19 - 1.86) |
| Non-Hospitalized | 3070 | 413 (13.5) | 0.91 (0.83 - 1.00) | 0.95 (0.86 - 1.04) | 211 | 24 (11.4) | 0.91 (0.61 - 1.36) | 1.04 (0.66 - 1.65) | 943 | 124 (13.1) | 0.85 (0.72 - 1.01) | 0.85 (0.72 - 1.01) | 2 | 265 (13.8) | 0.91 (0.81 - 1.03) | 1.02 (0.91 - 1.15) |
| Hospitalized | 65 | 15 (23.1) | 1.75 (1.16 - 2.63) | 1.13 (0.74 - 1.75) | 1 | 0 (0) | - | - | 12 | 1 (8.3) | 0.91 (0.15 - 5.49) | 0.51 (0.07 - 3.87) | 52 | 14 (26.9) | 1.74 (1.14 - 2.65) | 1.24 (0.80 - 1.91) |
| **COVID-19 related distress symptoms (PC-PTSD-5 / PCL-5)** | | | | | | | | | | | | | | | | |
| No confirmed diagnosis | 20 134 | 10 702 (53.2) | Ref. | Ref. | - | - | - | - | - | - | - | - | - | - | - | - |
| Ever diagnosed with COVID-19 | 2800 | 1546 (55.2) | 1.03 (0.99 - 1.07) | 1.03 (1.00 - 1.07) | 205 | 118 (57.6) | 1.08 (0.95 - 1.23) | 1.15 (0.97 - 1.36) | 883 | 483 (54.7) | 1.03 (0.97 - 1.10) | 1.03 (0.97 - 1.10) | 1712 | 945 (55.2) | 1.02 (0.98 - 1.07) | 1.04 (1.00 - 1.09) |
| Never bedridden | 456 | 216 (47.4) | 0.89 (0.81 - 0.98) | 0.92 (0.84 - 1.02) | 12 | 7 (58.3) | 1.08 (0.68 - 1.72) | 1.07 (0.67 - 1.72) | 208 | 98 (47.1) | 0.89 (0.77 - 1.03) | 0.91 (0.79 - 1.04) | 236 | 111 (47.0) | 0.88 (0.77 - 1.01) | 0.94 (0.82 - 1.07) |
| Bedridden 1-6 days | 432 | 229 (53.0) | 0.98 (0.90 - 1.07) | 0.99 (0.91 - 1.08) | 16 | 8 (50.0) | 0.99 (0.61 - 1.61) | 0.99 (0.60 - 1.63) | 183 | 98 (53.6) | 1.00 (0.88 - 1.15) | 1.00 (0.87 - 1.14) | 233 | 123 (52.8) | 0.97 (0.86 - 1.10) | 0.99 (0.87 - 1.12) |
| Bedridden 7 days or more | 366 | 241 (65.8) | 1.22 (1.13 - 1.31) | 1.15 (1.07 - 1.24) | 9 | 5 (55.6) | 1.06 (0.60 - 1.88) | 0.94 (0.53 - 1.69) | 98 | 61 (62.2) | 1.19 (1.03 - 1.39) | 1.12 (0.96 - 1.31) | 259 | 175 (67.6) | 1.23 (1.13 - 1.34) | 1.18 (1.08 - 1.28) |

| | | | | | | | | | | | | | | | | |
|---|---|---|---|---|---|---|---|---|---|---|---|---|---|---|---|---|
| Non-Hospitalized | 2718 | 1494 (55.0) | 1.02 (0.99 - 1.06) | 1.03 (1.00 - 1.07) | 203 | 117 (57.6) | 1.08 (0.94 - 1.23) | 1.15 (0.97 - 1.36) | 868 | 474 (54.6) | 1.03 (0.96 - 1.09) | 1.03 (0.97 - 1.10) | 1647 | 903 (54.8) | 1.01 (0.97 - 1.06) | 1.04 (0.99 - 1.09) |
| Hospitalized | 63 | 38 (60.3) | 1.25 (1.06 - 1.47) | 1.11 (0.94 - 1.32) | 2 | 1 (50.0) | 1.28 (0.37 - 4.38) | 1.03 (0.38 - 2.78) | 12 | 9 (75.0) | 1.38 (0.89 - 2.15) | 1.17 (0.73 - 1.90) | 49 | 31 (63.3) | 1.23 (1.03 - 1.47) | 1.11 (0.93 - 1.34) |
| **Sleep quality (PSQI, EST-Q2, Single-item measures)** | | | | | | | | | | | | | | | | |
| No confirmed diagnosis | 20 508 | 4859 (23.7) | Ref. | Ref. | - | - | - | - | - | - | - | - | - | - | - | - |
| Ever diagnosed with COVID-19 | 2891 | 779 (26.9) | 1.10 (1.03 - 1.18) | 1.11 (1.04 - 1.19) | 206 | 56 (27.2) | 1.11 (0.86 - 1.42) | 1.08 (0.80 - 1.47) | 909 | 248 (27.3) | 1.14 (1.02 - 1.28) | 1.14 (1.01 - 1.27) | 1776 | 475 (26.7) | 1.07 (0.99 - 1.17) | 1.11 (1.02 - 1.21) |
| Never bedridden | 474 | 98 (20.7) | 0.83 (0.69 - 0.99) | 0.89 (0.74 - 1.06) | 12 | 1 (8.3) | 0.33 (0.05 - 2.11) | 0.32 (0.06 - 1.84) | 211 | 42 (19.9) | 0.82 (0.63 - 1.08) | 0.84 (0.64 - 1.11) | 251 | 55 (21.9) | 0.86 (0.68 - 1.09) | 0.97 (0.76 - 1.22) |
| Bedridden 1-6 days | 461 | 130 (28.2) | 1.14 (0.98 - 1.32) | 1.15 (0.99 - 1.33) | 16 | 4 (25.0) | 1.06 (0.45 - 2.49) | 1.04 (0.42 - 2.54) | 193 | 61 (31.6) | 1.32 (1.07 - 1.62) | 1.31 (1.06 - 1.62) | 252 | 65 (25.8) | 1.01 (0.82 - 1.25) | 1.04 (0.84 - 1.29) |
| Bedridden 7 days or more | 380 | 142 (37.4) | 1.56 (1.37 - 1.78) | 1.40 (1.22 - 1.59) | 9 | 4 (44.4) | 1.83 (0.89 - 3.77) | 1.52 (0.81 - 2.87) | 100 | 36 (36.0) | 1.57 (1.20 - 2.05) | 1.40 (1.07 - 1.83) | 271 | 102 (37.6) | 1.52 (1.30 - 1.78) | 1.39 (1.19 - 1.62) |
| Non-Hospitalized | 2806 | 740 (26.4) | 1.07 (1.00 - 1.15) | 1.09 (1.02 - 1.16) | 204 | 55 (27.0) | 1.09 (0.85 - 1.41) | 1.07 (0.78 - 1.45) | 894 | 239 (26.7) | 1.11 (0.99 - 1.25) | 1.11 (0.99 - 1.25) | 1708 | 446 (26.1) | 1.04 (0.95 - 1.13) | 1.09 (1.00 - 1.19) |
| Hospitalized | 66 | 32 (48.5) | 2.05 (1.62 - 2.60) | 1.62 (1.27 - 2.08) | 2 | 1 (50.0) | 2.67 (0.66 - 10.74) | 1.89 (0.66 - 5.46) | 12 | 7 (58.0) | 3.02 (1.96 - 4.65) | 2.38 (1.68 - 3.36) | 52 | 24 (46.2) | 1.80 (1.36 - 2.39) | 1.47 (1.09 - 2.00) |

| | **CovidLife (UK)** N=18 518 | | | | | | | | | | | | | | | |
|---|---|---|---|---|---|---|---|---|---|---|---|---|---|---|---|---|
| | | **Overall** | | | | **Diagnosed 0-2 months ago** | | | | **Diagnosed 2-6 months ago** | | | | **Diagnosed 6-16 months ago** | | |
| | **n** | **$n_{threshold}$(%)** | **PR (age+gender adj)** | **PR (multivar adj)** [a] | **n** | **$n_{threshold}$(%)** | **PR (age+gender adj)** | **PR (multivar adj)** [a] | **n** | **$n_{threshold}$(%)** | **PR (age+gender adj)** | **PR (multivar adj)** [a] | **n** | **$n_{threshold}$ (%)** | **PR (age+gender adj)** | **PR (multivar adj)** [a] |
| **Depression (PHQ-9 / EST-Q2)** | | | | | | | | | | | | | | | | |
| No confirmed diagnosis | 17 492 | 3592 (20.5) | Ref. | Ref. | - | - | - | - | - | - | - | - | - | - | - | - |

| | | | | | | | | | | | | | | | | |
|---|---|---|---|---|---|---|---|---|---|---|---|---|---|---|---|---|
| Ever diagnosed with COVID-19 | 312 | 100 (32.1) | 1.41 (1.19-1.67) | 1.34 (0.96-1.89) | 205 | 71 (34.6) | 1.50 (1.23-1.83) | 1.35 (0.96-1.90) | 97 | 26 (26.8) | 1.20 (0.85-1.69) | 1.36 (0.99-1.87) | 10 | 3 (30) | 1.40 (0.56-3.51) | 1.74 (0.59-5.17) |
| Never bedridden | - | - | - | - | - | - | - | - | - | - | - | - | - | - | - | - |
| Bedridden 1-6 days | - | - | - | - | - | - | - | - | - | - | - | - | - | - | - | - |
| Bedridden 7 days or more | - | - | - | - | - | - | - | - | - | - | - | - | - | - | - | - |
| Non-Hospitalized | 275 | 80 (29.1) | 1.27 (1.05-1.55) | 1.06 (0.70-1.60) | 173 | 53 (30.6) | 1.30 (1.02-1.66) | 0.98 (0.62-1.54) | 92 | 24 (26.1) | 1.21 (0.85-1.70) | 1.39 (1.01-1.92) | 10 | 3 (30) | 1.40 (0.56-3.51) | 1.74 (0.59-5.17) |
| Hospitalized | 34 | 17 (50) | 2.39 (1.68-3.39) | 1.96 (1.39-2.76) | 29 | 15 (51.7) | 2.59 (1.86-3.60) | 2.15 (1.55-2.98) | 5 | 2 (40) | 1.10 (0.15-8.31) | 0.94 (0.20-4.43) | - | - | - | - |
| **Anxiety symptoms (GAD-7 / ASS / EST-Q2)** | | | | | | | | | | | | | | | | |
| No confirmed diagnosis | 17 533 | 2997 (17.1) | Ref. | Ref. | - | - | - | - | - | - | - | - | - | - | - | - |
| Ever diagnosed with COVID-19 | 314 | 70 (22.3) | 1.18 (0.96-1.46) | 1.37 (0.92-2.04) | 207 | 51 (24.6) | 1.26 (0.99-1.62) | 1.38 (0.93-2.05) | 97 | 16 (16.5) | 0.96 (0.62-1.47) | 1.07 (0.70-1.65) | 10 | 3 (30) | 1.68 (0.56-5.09) | 2.00 (0.67-5.97) |
| Never bedridden | - | - | - | - | - | - | - | - | - | - | - | - | - | - | - | - |
| Bedridden 1-6 days | - | - | - | - | - | - | - | - | - | - | - | - | - | - | - | - |
| Bedridden 7 days or more | - | - | - | - | - | - | - | - | - | - | - | - | - | - | - | - |
| Non-Hospitalized | 277 | 56 (20.2) | 1.07 (0.84-1.36) | 1.14 (0.70-1.84) | 175 | 37 (21.1) | 1.06 (0.78-1.44) | 1.05 (0.62-1.78) | 92 | 16 (17.4) | 1.00 (0.65-1.54) | 1.13 (0.74-1.74) | 10 | 3 (30) | 1.68 (0.56-5.09) | 2.00 (0.67-5.97) |
| Hospitalized | 34 | 11 (32.4) | 1.97 (1.26-3.07) | 1.91 (1.18-3.08) | 29 | 11 (37.9) | 2.29 (1.51-3.46) | 2.13 (1.34-3.39) | 5 | 0 (0) | - | - | - | - | - | - |
| **COVID-19 related distress symptoms (PC-PTSD-5 / PCL-5)** | | | | | | | | | | | | | | | | |
| No confirmed diagnosis | - | - | - | - | - | - | - | - | - | - | - | - | - | - | - | - |
| Ever diagnosed with COVID-19 | - | - | - | - | - | - | - | - | - | - | - | - | - | - | - | - |
| Never bedridden | - | - | - | - | - | - | - | - | - | - | - | - | - | - | - | - |

| | | | | | | | | | | | | | | | | |
|---|---|---|---|---|---|---|---|---|---|---|---|---|---|---|---|---|
| Bedridden 1-6 days | - | - | - | - | - | - | - | - | - | - | - | - | - | - | - | - |
| Bedridden 7 days or more | - | - | - | - | - | - | - | - | - | - | - | - | - | - | - | - |
| Non-Hospitalized | - | - | - | - | - | - | - | - | - | - | - | - | - | - | - | - |
| Hospitalized | - | - | - | - | - | - | - | - | - | - | - | - | - | - | - | - |
| **Sleep quality (PSQI, EST-Q2, Single-item measures)** | | | | | | | | | | | | | | | | |
| No confirmed diagnosis | 17 374 | 5155 (29.7) | Ref. | Ref. | - | - | - | - | - | - | - | - | - | - | - | - |
| Ever diagnosed with COVID-19 | 293 | 108 (36.9) | 1.12 (0.96-1.32) | 1.03 (0.74-1.44) | 190 | 67 (35.3) | 1.08 (0.88-1.32) | 1.04 (0.75-1.45) | 93 | 36 (38.7) | 1.20 (0.92-1.57) | 1.23 (0.94-1.62) | 10 | 5 (50) | 1.33 (0.61-2.87) | 1.39 (0.62-3.13) |
| Never bedridden | - | - | - | - | - | - | - | - | - | - | - | - | - | - | - | - |
| Bedridden 1-6 days | - | - | - | - | - | - | - | - | - | - | - | - | - | - | - | - |
| Bedridden 7 days or more | - | - | - | - | - | - | - | - | - | - | - | - | - | - | - | - |
| Non-Hospitalized | 260 | 92 (35.4) | 1.08 (0.90-1.28) | 0.94 (0.64-1.37) | 162 | 52 (32.1) | 0.96 (0.75-1.22) | 0.84 (0.55-1.28) | 88 | 35 (39.8) | 1.26 (0.97-1.64) | 1.31 (1.00-1.71) | 10 | 5 (50) | 1.33 (0.61-2.87) | 1.39 (0.62-3.13) |
| Hospitalized | 31 | 14 (45.2) | 1.43 (0.95-2.16) | 1.28 (0.82-2.01) | 26 | 13 (50) | 1.67 (1.14-2.45) | 1.55 (1.03-2.35) | 5 | 1 (20) | 0.00 (0.00-0.00) | 0.00 (0.00-0.00) | - | - | - | - |

| | Overall<br>N = 247 519 | | | | | | | |
|---|---|---|---|---|---|---|---|---|
| | **Overall** | | **Diagnosed 0-2 months ago** | | **Diagnosed 2-6 months ago** | | **Diagnosed 6-16 months ago** | |
| | **n** | **$n_{threshold}$ (%)** | **n** | **$n_{threshold}$ (%)** | **n** | **$n_{threshold}$ (%)** | **n** | **$n_{threshold}$ (%)** |
| **Depression (PHQ-9 / EST-Q2)** | | | | | | | | |
| No confirmed diagnosis | 197243 | 22241 (11.3) | - | | | | | |
| Ever diagnosed with COVID-19 | 8150 | 1643 (20.2) | 2547 | 523 (20.5) | 3192 | 611 (19.1) | 2393 | 507 (21.2) |
| Never bedridden | 2585 | 353 (13.7) | 1001 | 136 (13.6) | 1163 | 155 (13.3) | 421 | 65 (15.4) |
| Bedridden 1-6 days | 2230 | 452 (20.3) | 866 | 189 (21.8) | 961 | 197 (20.5) | 403 | 63 (15.6) |
| Bedridden 7 days or more | 1030 | 299 (29) | 272 | 77 (28.3) | 438 | 112 (25.6) | 320 | 110 (34.4) |

| | | | | | | | | |
|---|---|---|---|---|---|---|---|---|
| Non-Hospitalized | 6593 | 1373 (20.8) | 1840 | 400 (21.7) | 2645 | 529 (20) | 2108 | 444 (21.1) |
| Hospitalized | 200 | 69 (34.5) | 64 | 27 (42.2) | 70 | 13 (18.6) | 66 | 29 (43.9) |
| **Anxiety symptoms (GAD-7 / ASS / EST-Q2)** | | | | | | | | |
| No confirmed diagnosis | 204151 | 15140 (7.4) | | | | | | |
| Ever diagnosed with COVID-19 | 8249 | 1076 (13) | 2631 | 362 (13.8) | 3159 | 393 (12.4) | 2459 | 317 (12.9) |
| Never bedridden | 2602 | 267 (10.3) | 1005 | 118 (11.7) | 1159 | 107 (9.2) | 438 | 38 (8.7) |
| Bedridden 1-6 days | 2257 | 300 (13.3) | 868 | 122 (14.1) | 969 | 135 (13.9) | 420 | 40 (9.5) |
| Bedridden 7 days or more | 1047 | 194 (18.5) | 283 | 48 (17) | 436 | 77 (17.7) | 328 | 69 (21) |
| Non-Hospitalized | 6688 | 971 (14.5) | 1856 | 313 (16.9) | 2660 | 366 (13.8) | 257.916 | 292 (113.2) |
| Hospitalized | 203 | 39 (19.2) | 68 | 18 (26.5) | 67 | 6 (9) | 68 | 15 (22.1) |
| **COVID-19 related distress symptoms (PC-PTSD-5 / PCL-5)** | | | | | | | | |
| No confirmed diagnosis | 74902 | 23859 (31.9) | | | | | | |
| Ever diagnosed with COVID-19 | 5165 | 2116 (41) | 1260 | 401 (31.8) | 1840 | 713 (38.8) | 2065 | 1002 (48.5) |
| Never bedridden | 1444 | 416 (28.8) | 482 | 113 (23.4) | 603 | 175 (29) | 359 | 128 (35.7) |
| Bedridden 1-6 days | 1438 | 505 (35.1) | 464 | 140 (30.2) | 603 | 214 (35.5) | 371 | 151 (40.7) |
| Bedridden 7 days or more | 627 | 316 (50.4) | 129 | 47 (36.4) | 197 | 90 (45.7) | 301 | 179 (59.5) |
| Non-Hospitalized | 3774 | 1693 (44.9) | 610 | 213 (34.9) | 1373 | 568 (41.4) | 1791 | 912 (50.9) |
| Hospitalized | 124 | 55 (44.4) | 24 | 10 (41.7) | 42 | 16 (38.1) | 58 | 32 (55.2) |
| **Sleep quality (PSQI, BIS, EST-Q2, Single-item measures)** | | | | | | | | |
| No confirmed diagnosis | 143265 | 34060 (23.8) | | | | | | |
| Ever diagnosed with COVID-19 | 7510 | 2211 (29.4) | 2100 | 620 (29.5) | 3065 | 947 (30.9) | 2345 | 644 (27.5) |
| Never bedridden | 2361 | 624 (26.4) | 746 | 198 (26.5) | 1144 | 308 (26.9) | 471 | 118 (25.1) |
| Bedridden 1-6 days | 1813 | 538 (29.7) | 630 | 182 (28.9) | 829 | 265 (32) | 354 | 91 (25.7) |
| Bedridden 7 days or more | 1338 | 499 (37.3) | 329 | 117 (35.6) | 545 | 217 (39.8) | 464 | 165 (35.6) |
| Non-Hospitalized | 7107 | 2065 (29.1) | 1981 | 575 (29) | 2928 | 898 (30.7) | 2198 | 592 (26.9) |
| Hospitalized | 213 | 92 (43.2) | 63 | 29 (46) | 72 | 29 (40.3) | 78 | 34 (43.6) |

[a] Adjusted for gender, age, education, relationship status, smoking, BMI, previous psychiatric diagnosis, number of comorbidities and response period

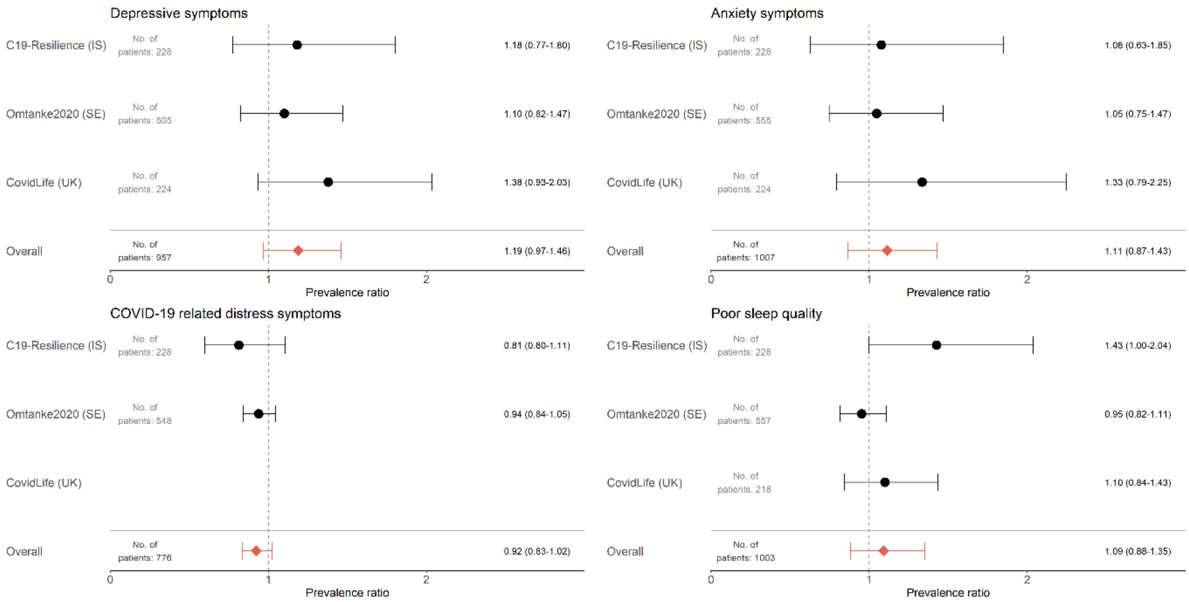

Figure S2: **Change in mental health indicators before and after diagnosis of COVID-19, sub-analysis within cohorts with repeated measures (Median time between follow up for cohorts: C19-Resilience: 7.3 months, Omtanke2020: 1 month, CovidLife: 10 months).**

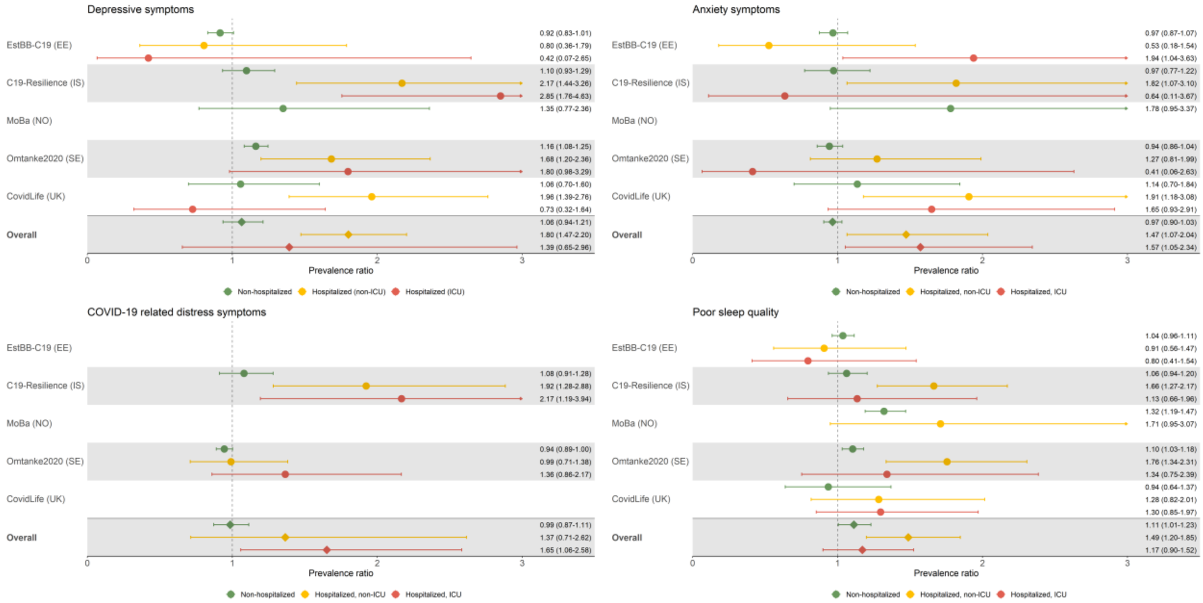

Figure S3: **Mental health indicators among individuals with a diagnosis of COVID-19 compared with individuals without COVID-19 by hospitalization status.**

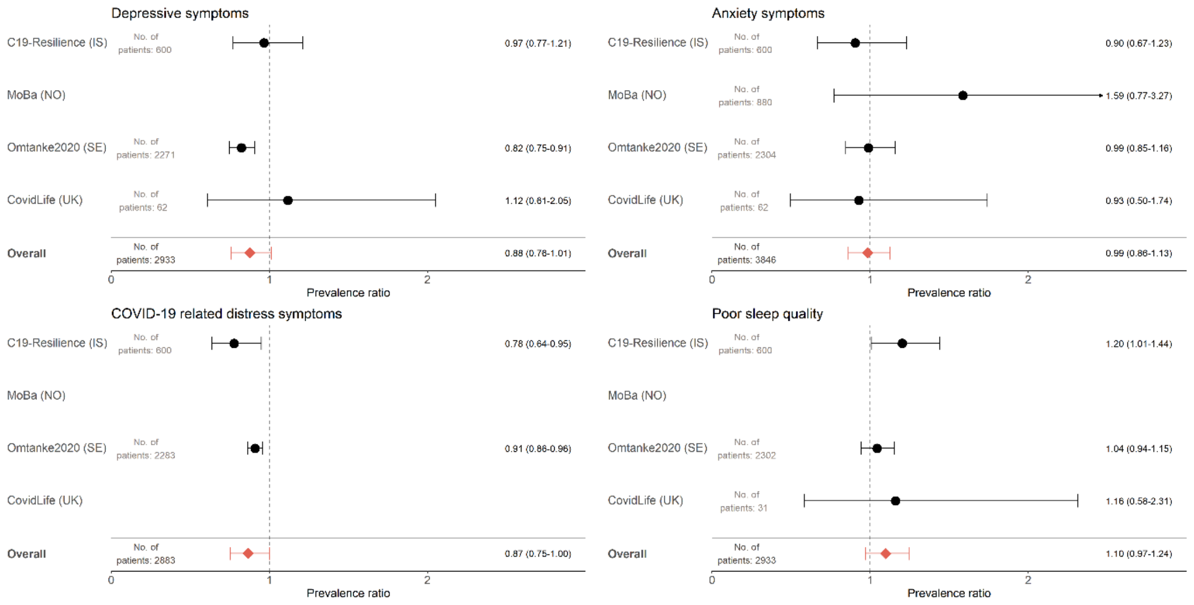

Figure S4: **Change in mental health indicators from first to last measure after diagnosis of COVID-19, sub-analysis of patients with COVID-19 with repeated measures (Median time between follow up for cohorts: C19-Resilience: 12.7 months, MoBa: 10.1 months, Omtanke2020: 6 months, CovidLife: 10 months).**

Table S5: **Overall prevalence ratios of mental health indicators during the first 16 months after diagnosis of COVID-19 by illness severity**

| Mental health indicator | Time since diagnosis | Time spent bedridden | PR (95% confidence interval) |
|---|---|---|---|
| *Depressive symptoms* | 0–2 months | Never bedridden | 0·84 (0·69-1·03) |
| | | 1–6 days | 1·32 (0·85-2·04) |
| | | > 7 days | 1·66 (1·11-2·47) |
| | 2–6 months | Never bedridden | 0·76 (0·66-0·89) |
| | | 1–6 days | 1·21 (0·89-1·63) |
| | | > 7 days | 1·53 (1·26-1·86) |
| | 6–16 months | Never bedridden | 0·96 (0·77-1·19) |
| | | 1–6 days | 0·97 (0·78-1·21) |
| | | > 7 days | 1·60 (1·17-2·18) |
| *Anxiety symptoms* | 0–2 months | Never bedridden | 0·91 (0·78-1·07) |
| | | 1–6 days | 1·01 (0·86-1·18) |
| | | > 7 days | 1·47 (0·80-2·72) |
| | 2–6 months | Never bedridden | 0·71 (0·55-0·92) |
| | | 1–6 days | 1·03 (0·87-1·22) |
| | | > 7 days | 1·46 (1·20-1·79) |
| | 6–16 months | Never bedridden | 0·87 (0·60-1·26) |
| | | 1–6 days | 0·82 (0·58-1·17) |
| | | > 7 days | 1·47 (1·19-1·81) |

| | | | |
|---|---|---|---|
| *COVID-19 related distress symptoms* | 0−2 months | Never bedridden | 1·01 (0·86-1·19) |
| | | 1−6 days | 1·22 (1·06-1·39) |
| | | > 7 days | 1·78 (1·29-2·46) |
| | 2−6 months | Never bedridden | 0·82 (0·47-1·42) |
| | | 1−6 days | 1·05 (0·86-1·27) |
| | | > 7 days | 1·17 (0·93-1·47) |
| | 6−16 months | Never bedridden | 0·88 (0·73-1·06) |
| | | 1−6 days | 0·91 (0·72-1·15) |
| | | > 7 days | 1·01 (0·52-1·96) |
| *Poor sleep quality* | 0−2 months | Never bedridden | 0·94 (0·84-1·06) |
| | | 1−6 days | 0·98 (0·86-1·11) |
| | | > 7 days | 1·35 (1·11-1·64) |
| | 2−6 months | Never bedridden | 0·98 (0·87-1·11) |
| | | 1−6 days | 1·14 (0·96-1·35) |
| | | > 7 days | 1·48 (1·29-1·69) |
| | 6−16 months | Never bedridden | 1·02 (0·87-1·20) |
| | | 1−6 days | 0·95 (0·71-1·27) |
| | | > 7 days | 1·26 (0·90-1·77) |
